# Exploring Motor Imagery as a Therapeutic Intervention for Parkinson’s Disease Patients: A Scoping Review

**DOI:** 10.1101/2024.03.29.24305071

**Authors:** Maxime Michel, Elena Terragno, Matthieu Bereau, Eloi Magnin, Nicolas Gueugneau, Antonio Vinicius Soares, Yoshimasa Sagawa

## Abstract

**Background:** Motor Imagery (MI) has emerged as a promising therapeutic approach in the rehabilitation of individuals with Parkinson’s Disease (PD). MI entails mentally rehearsing motor actions without physically executing them. This cognitive process has garnered attention due to its potential benefits in aiding motor function recovery in PD patients. Its role in complementing traditional treatment approaches is likely to reverberate throughout clinical practice. This study strives to provide a comprehensive examination several MI protocols designed for individuals with PD. The focus was to underscore the outcomes observed across motor symptoms, balance, gait, and quality of life.

**Methods:** A literature search was carried out in the following databases: Medline, Embase, Cochrane, and PEDro, from the first publication to February 2024. Study with at least one keyword in relation to PD and MI in the title were included.

**Results:** Of the 262 studies 53 were included. Twelve RCTs with a mean PEDro score of 6.6/10 and 41 descriptive and non-RCT studies. Among the RCTs, there were almost exclusively MI on balance, gait, and lower limbs exercise. They found an 85.2% improvement for the experimental group on the TUG with a cognitive task (p<0.02), 5.8% on the TUG (p<0.05), a 5.1% improvement in walking speed (p<0.05), other variables did not show significant improvement. For the descriptive and non-RCTs studies, there were various tasks and outcomes for the lower and upper limbs. It was shown that there was no difference in execution time in MI between patients with PD and HS, while in ME patients with PD were slower. For the upper limb, several tasks were proposed, such as thumb opposition, joystick movements and writing tasks with variable results. RCTs were more focused on balance, lower limb and walking, there was no specific outcome for the upper limb and speech. The heterogeneity of the tasks and outcomes across all included studies is also a limitation.

**Conclusion:** To summarize, the current research on walking disorders in PD shows promise, but further investigations are crucial, particularly with an emphasis on upper limb function and speech. A need exists for studies with larger sample sizes, utilizing precise methodologies, and specifically targeting these areas to enhance our comprehension of the potential advantages of MI in the context of comprehensive PD rehabilitation.

## 1 Introduction

Parkinson’s disease (PD) is the second most common neurodegenerative disorder after Alzheimer’s disease and a major cause of disability in the elderly. The prevalence of patients with PD is expected to double between 2015 and 2030, particularly due to the aging of the population (1). Indeed, age is the main risk factor for this pathology (2). PD is caused by loss of dopaminergic neurons and causes motor and non-motor symptoms (2,3). The three notable motors symptoms are akinesia, rest tremor and rigidity (2–10) whereas the non-motor symptoms include sleep disorders, depression, and digestive disorders (11). PD affects sensorimotor functions as walking, balance, posture and have a negative impact on patient’s independency and societal participation (12).

Different treatments exist in PD, such as pharmacological treatments based on dopamine and its derived which is the most common one (4). While levodopa is widely recognised as the most effective medication for treating the motor symptoms, there are other medications such as monoamine oxidase type B inhibitors, amantadine, anticholinergics, β-blockers, or dopamine agonists. Its utilisation is conditioned by the symptoms expressed by the patient (13). Although this treatment is the most used, side effects as dyskinesias and motor complications can be observed (14). This is one of the main reasons as other forms of symptomatic treatment were researched. Among non-pharmacological treatments, physiotherapy has shown beneficial effects in the management of PD (5). Recent studies have been showing positive effects on motor symptoms (5), quality of life (15), walking and balance (5,16,17).

Among the physiotherapy’s techniques, motor imagery (MI) was proposed more than 30 years ago as a potential tool of rehabilitation (18). It is defined as a mental process where a person make a mental simulation of a motor act without making any movement (7,8). This approach relies on the premise that MI and actual motor execution elicit activation in overlapping brain areas (19). Consequently, enhancing the engagement of motor regions in the brain (9) is a central objective of this technique.

The MI, a recently developed approach for the rehabilitation of patients with PD, is supported and promoted for implementation in rehabilitation protocols as a promising approach (6,20,21). Some studies have demonstrated the effectiveness of MI combined to physiotherapy on patients with PD (6,22). The MI can be performed from first person or a third person’s perspective (7,23), and can be used for different modalities such as upper limb, lower limb, walking, and others. Also, there are numerous protocols with distinct sensorimotor tasks. (24–29): e.g. goal-directed task and Box and Block Test (BBT) (26), MI of walking along a straight course (24), MI of walking forward, backward and turning (25). Considering these different modalities of MI, the choice of the best MI protocol for a clinical application seems difficult; how a MI protocol should be conducted and how benefits should be expected. Only one study proposes a framework for motivational interviewing MI aimed at aiding physiotherapists in integrating MI into their clinical practice (27). Aligned with the imperative to optimize the clinical use of MI as a rehabilitation tool MI, this scoping review sought to achieve two primary objectives. Firstly, it aimed to provide a comprehensive summary of the diverse protocols of MI designed for patients with Parkinson’s Disease (PD), with the goal of offering guidance and facilitating their application in clinical practice. Secondly, the review aimed to highlight the key findings observed in these studies concerning motor symptoms, balance, gait, and quality of life.

## 2 Materials and Methods

This review was conducted in accordance with the Preferred Reporting Items for Systematic review and Meta-Analyses extension for Scoping Reviews (PRISMA-ScR) guidelines (Annex I). According with our previous research, to date, there is no scoping review existing on this subject.

### 2.1 Data sources and searches

Prospective research was carried out on four different databases: MEDLINE (PubMed), Embase, Cochrane (Cochrane library) and PEDro from the first publication until February 2024. To select relevant articles, the follow keywords and operators were used: “Parkinson disease”* OR “Parkinson Disease” OR “Parkinson’s disease”* AND “motor imagery”* OR “motor imagery practice”* OR “mental practice”*. To improve exhaustiveness of potential articles included, the search was conducted with Medical Subject Heading (MeSH) terms and non-MeSH terms (identified by an asterisk).

### 2.2 Study selection

Firstly, all articles with at least one keyword in relation to PD and MI in the title were included in this phase. Duplicated articles were removed. The eligibility criteria (Figure 1) for this phase of selection were applied to titles and abstracts of the articles. Exclusion criteria were articles that are neither in English nor in French, feasibility and pilot study, conference abstract, articles that do not focus on the specific effectiveness of MI. Full text was directly reviewed with eligibility criteria when abstract did not provide sufficient information. Then, eligibility criteria were applied to full text.

**Figure 1.**
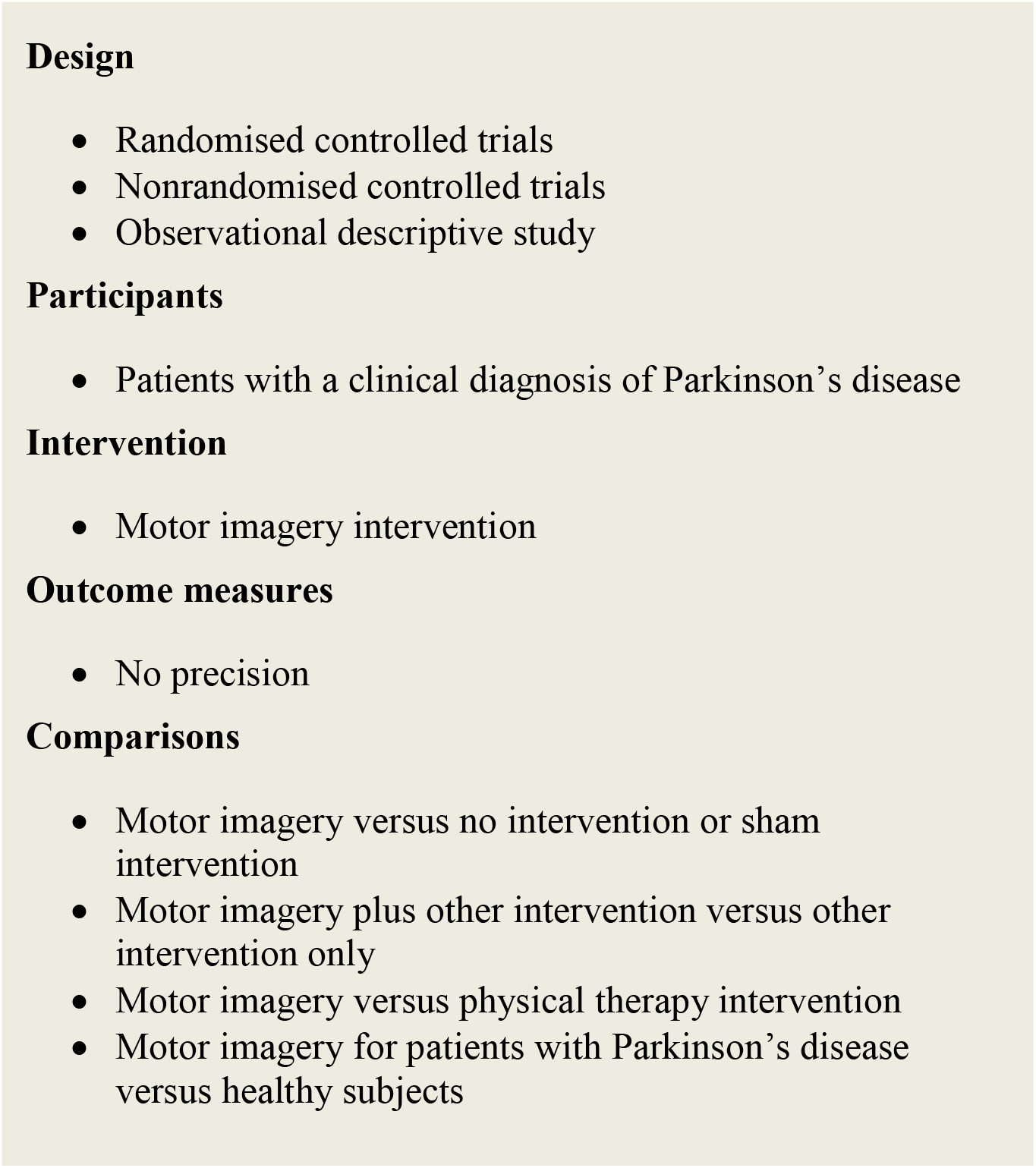
Eligibility criteria

### 2.3 Data extraction and quality assessment

For this review, articles were selected and read by two reviewers (MM and ET). Disagreements in this phase were resolved by consulting a third evaluator (YS). Methodological quality of the randomised controlled studies (RCTs) was assessed with PEDro scale. This is an 11-item scale. It is used to assessed external validity (criterion 1), internal validity (criterion 2 to 9) and interpretability of the findings (criterion 10 and 11) of a clinical trial or group comparison study. The PEDro scale scored in 10 points (0 very poor methodological quality 10 excellent methodological quality).

### 2.4 Data synthesis and analysis

Reviewers extracted the following key data for each article: type of study, population characteristics, inclusion/exclusion criteria, intervention/protocol, variable of interest and PEDro score. Mean (±SD) values for all variables, *p* values and modification in percentage (comparisons among interventions, groups) were collected.

## 3 RESULTS AND COMMENTS

### 3.1 Selection of articles

Figure 2 presents the article selection process of this review. From the 4 databases combined, 262 articles were identified. Fifty-three of these articles were included, with 12 RCTs and 41 non-RCTs and descriptive studies. Methodological quality as assessed by the mean PEDro score for RCTs was 6.6/10, only one was lowest than 3/10 (30). Eligibility criteria, random allocation, baseline intragroup similarity and between group statistical comparison were respected for all studies. Subjects and therapists blinding were not respected for all RCTs.

**Figure 2.**
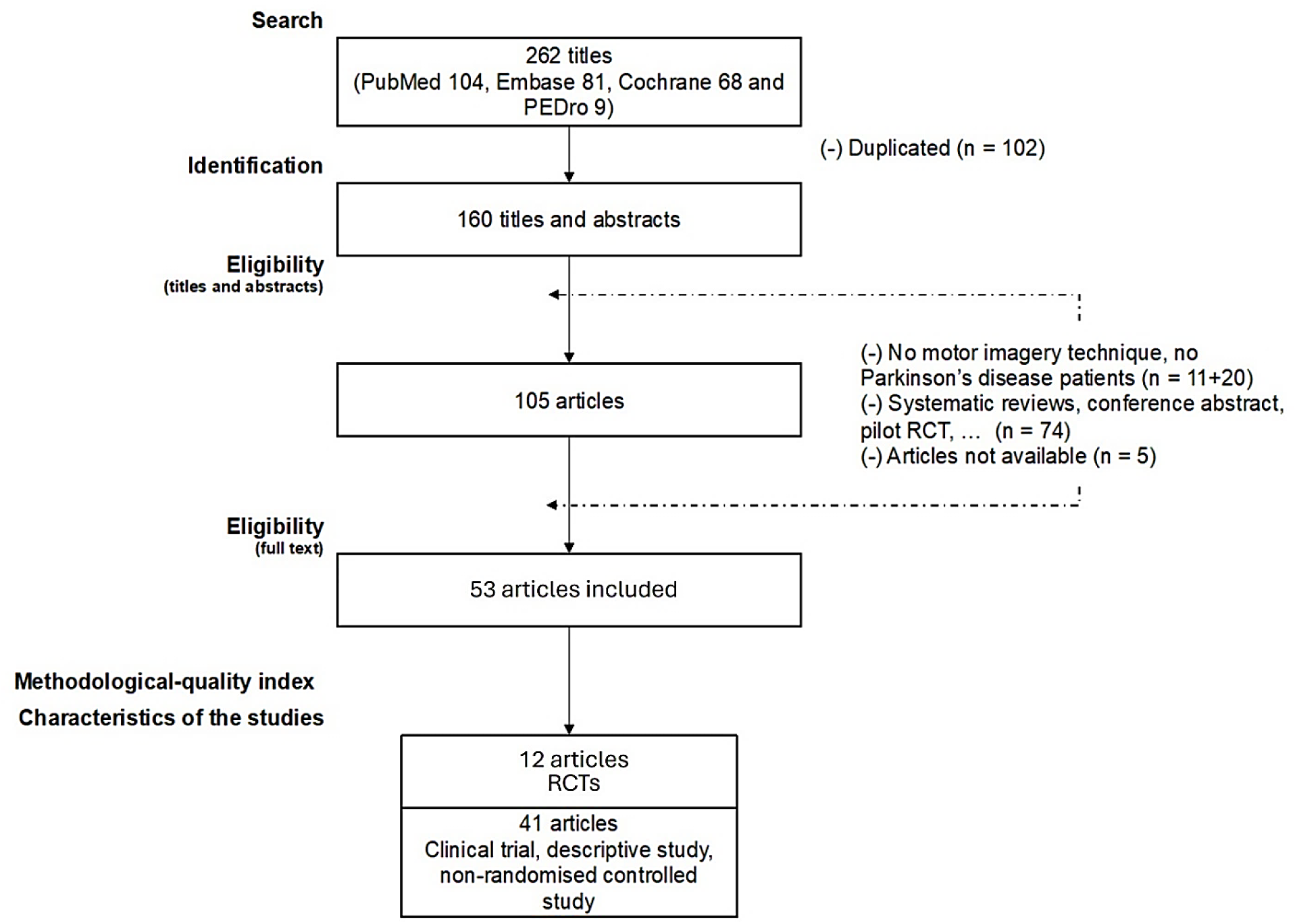
Flow of studies for the review

### 3.2 RCT: effects of MI intervention

#### 3.2.1 Participants characteristics

The characteristics of RCTs are presented in Table 1. Participant’s characteristics were based on diagnosis of PD. The mean (SD) number of participants per study was 29.9 (±10.5) with a mean age of 66.2 (±8.3) years old. Groups were composed on average of 30.7% women and 69.3% men. Mean (SD) Hoehn and Yahr (H&Y) score was 2.2 (0.5) with the off-phase score taken when it was specified.

**Table 1.** Characteristics of the randomised controlled trials.

| Articles | Type of study | Participants: nb (nb per gender), mean (SD) age, mean (SD) UPDRS stage, mean (SD) H&Y score, treatment | Inclusion criteria (diagnosis, age, H&Y scale, MMSE score, others) | Exclusion criteria | Protocol (task, sessions [No. and W], frequency, intensity) | Evaluation (No., date, and outcomes) | PEDro score |
| --- | --- | --- | --- | --- | --- | --- | --- |
| Sarasso et al. (2023) | RCT | Experimental group: 10 PD patients (5♀), 67.6 (6.4) y, H&Y OFF 5/4/3, UPDRS III 33.1 (11.9)<br>Control group: 12 PD patients (5♀), 64.1 (8.9) y, H&Y OFF 5/4/4, UPDRS III 33.8 (10.5) | Idiopathic PD, H&Y score ≤3. Mini-mental score examination (MMSE) score (greater than or equal to 24) | Medical illnesses or substance abuse that could interfere with cognition; any (other) major systemic, psychiatric, neurological, visual, and musculoskeletal disturbances or other causes of walking inability; contraindications to undergoing MRI examination; and brain damage at routine MRI, including lacunae and extensive cerebrovascular disorders | Experimental group: Performed DUAL-TASK + AOT-MI (four gait/balance exercises each session were proposed with the following modality: 2 min of task observation → 5 min of task execution → 2 min of task imagination → 5 min of task execution) Control group: DUAL-TASK performed the same number of exercises combined with watching landscape videos instead of observation/imagination<br>For both groups: 1 h each session, 3 d/wk for 6 wks | Primary clinical outcome: Kinesthetic and Visual Imagery Questionnaire (KVIQ) version 10, and brain MRI scans | 7 |
| Bezerra et al. (2022) | RCT | Experimental group: 21 PD patients (7♀), 64.6 (9.3) y, H&Y OFF 2.0 (2.0-3.0), UPDRS II 23.0 (15.5-32.5), UPDRS III 13.0 (9.0-18.5)<br>Control group: 18 PD patients (7♀), 60.7 (6.8) y, H&Y OFF 2.5 (2.0-3.0), UPDRS II 27.5 (18.0-41.2), UPDRS III 14.0 (10.0-23.0) | Idiopathic PD, H&Y scores 1.5 to 3; regular use of antiparkinsonian medication; walk independently for at least 10 meters without any orthosis or gait aid; no cognitive deficit according to the Mini-Mental state Examination (cutoff of 18 points for illiterate and 24 for those with school education) | Musculoskeletal or cardiorespiratory impairments affecting gait; and absence of other associated neurological diseases | Experimental group: Performed 12 sessions of AO, MI, and gait training.<br>Control group: Watched PD-related educational videos and performed 12 sessions of gait training.<br>For both groups: 1 h each session, 3 d/wk for 4 wks | Primary clinical outcome: MiniBESTest: Mini Balance Evaluation Systems Test; FOG-Q: freezing of Gait Questionnaire. | 8 |
| Kashif et al. (2022) | RCT | Experimental group: 22 PD patients (9♀), 63.9 (4.6) y, H&Y OFF 2.1 (0.7), UPDRS II 22.0 (4.6), UPDRS III 32.5 (4.0)<br>Control group: 22 PD patients (10♀), 2.3 (4.6) y, H&Y OFF 2.6 (0.7), UPDRS II 21.5 (3.9), UPDRS III 31.9 (4.6) | Idiopathic PD, severity ranging from stage I to stage III on the modified H and Y scale, intact cognition according to their mini-mental score examination (MMSE) score (greater than or equal to 24) | Other neurological presentation, orthopaedic pathology, visual anomalies, cardiovascular issues, severe dyskinesia or “on-off” phases, a history of surgery for PD, a history of virtual games used for treatment in the last three months, and virtual game phobia | Experimental group: Physiotherapy + virtual reality (Nintendo Wii) + motor imagery Control group: Physiotherapy<br>60 min/d | Primary clinical outcome: MDS-UPDRS part II and III<br>Secondary clinical outcome: Balance confidence and balance | 7 |
|  |  |  |  |  | For both groups: 3 d/wk, for 12 wks with follow-up to 16 wks |  |  |
| Tinaz et al. 2022 | RCT | Experimental group: 22 PD patients (12♀), 66.2 (8.1) y, MDS-UPDRS III 32.3 (8.1), H&Y OFF 2.0 (0.2), NI<br>Control group: 22 PD patients (12♀), 65.7 (8.8) y, MDS-UPDRS III 34.5 (9.6), H&Y OFF 2.1 (0.3), NI | Idiopathic Parkinson's disease (according to UK Brain Bank criteria)<br>Age ≥ 40 y<br>Stable dopaminergic treatment during the study | H&Y scale >stage 3<br>Not fully independent<br>Neurological or psychiatric disorder<br>Medical condition that might affect central nervous system<br>History of alcohol or illicit drug abuse<br>Head injury resulting in loss of consciousness<br>MoCA < 21<br>Contraindications for MRI<br>Poor homework compliance (<50%) | Experimental group: neurofeedback kinaesthetic MI (walking, balance exercises, calisthenics)<br>Control group: visual imagery exercises<br>For both groups: 4W, every day<br>Tested in off-state | 2 at W0 and after training<br>Primary clinical outcome: MDS-UPDRS part III<br>Secondary clinical outcome: 2 min endurance walking, TUG, 5 times sit-to-stand, 360-degree turning, physical performance test<br>Primary imaging outcome: change in right insula-dmFC functional connectivity strength | 5 |
| Sarasso et al. 2021 | RCT | Experimental group: 13 (5♀), 67.5 (6.1) y, MDS-UPDRS II 10.38 (5.55), H&Y ON 2.33 (0.5)/OFF 2.44 (0.5), NI<br>Control group: 12 (4♀), 63.8 (9.2) y, MDS-UPDRS II 12.58 (5.14), H&Y ON 2.38 (0.5)/OFF 2.5 (0.5), NI | H&Y score ≤4<br>Postural instability and gait disorders phenotype<br>Stable dopaminergic medication for at least 4 weeks, w/out any changes during observation period<br>No dementia, MMSE ≥24<br>No significant head tremor | Medical illnesses or substance abuse that could interfere w/ cognition<br>Other major systemic, psychiatric, neurological, visual, and musculoskeletal disturbances or other causes of walking inability<br>Contraindications to undergoing MRI examination<br>Brain damage at routine MRI including lacunae and extensive cerebrovascular disorders | Experimental group: gait/balance training with dual task exercises added with AOT-MI therapy<br>Control group: gait/balance training with dual task exercises and watching landscapes<br>For both groups: 6W, 3/W, 1 hour<br>Tested in on-state | 3 at W0, W6 and W14<br>TUG with cognitive (primary outcome)<br>TUG<br>TUG with manual dual task<br>MiniBESTest<br>ABC scale<br>10MWT<br>PDQ-39<br>NFOG-Q | 7 |
| Mahmoud et al. 2018 | RCT | Experimental group: 15 (4♀), NI, levodopa medication<br>Control group: 15 (5♀), NI, levodopa medication | Idiopathic Parkinsonism with cognitive dysfunctions (confirmed with RehaCom)<br>Age: between 50 and 65 years<br>Modified H&Y scale: stage 1 to 3<br>Male and female<br>Disease duration from 3 to 5 years<br>Taking levodopa medication | Other symptoms of Parkinsonism<br>Modified H&Y scale: stage 4 to 5<br>Damaged eyesight who could not recognise objects on a computer screen | Experimental group: MI with cues, relaxation, and breathing exercises, sit to stand task and exercises in standing position and the control group task<br>Control group: mental cognitive exercises including memory recall, maths exercises, mental arithmetic, dual tasking<br>For both groups: 6W, 3/W, 1 hour<br>Tested in on-state | 2 at pre-training and post-training<br>Attention and concentration level (RehaCom assessment tool)<br>Reaction time (RehaCom assessment tool)<br>Figural memory level and missed pictures | 3 |
| Monteiro et al. 2018 | RCT | Experimental group: 7 (0♀), 64 (7) y, UPDRS NS, H&Y OFF 2 (1), treatment NI<br>Control group: 7 (2♀), 62 (12) y, UPDRS NS, H&Y OFF 2 (0.5), treatment NI<br>Initially 22 patients with PD received intervention but they were 8 follow-up losses | Age between 45 to 72 years<br>H&Y scale: stage 1 to 3<br>Both genders | Other neurological diseases<br>Decompensated systemic diseases<br>Reduced cognitive level<br>Unable to perform MI during KVIQ-20 | All patients before randomization: motor physiotherapy<br>Experimental group: MI practice of a step and home exercises with handbook<br>Control group: home exercises with handbook<br>All patients before randomization: 15 sessions of 40 minutes, 2/W<br>MI practice: 10 sessions of 5-10 min, 2/W<br>Home exercises with handbook: 12W, 3/W, 50 min<br>Tested in on-state | 3 at baseline (evaluation), after motor physiotherapy (reevaluation 1), after mental practice (reevaluation 2)<br>TUG<br>DGI<br>FES-I Brazil | 7 |
| Subramanian et al. 2016 | RCT | Experimental group 15 (1♀), 67 (9) y, MDS-UPDRS-MS 23.3 (9.4), H&Y 1.6 (0.6), levodopa and equivalent medication<br>Control group 15 (3♀), 63 (11) y, MDS-UPDRS-MS 26.7 (12.6), H&Y 1.7 (0.5), levodopa and equivalent medication | Diagnosis of PD<br>H&Y scale: stage 1 to 3<br>No dementia or significant comorbidity and fulfilled safety requirements for MRI | NI | Experimental group: Homework employing MI + supervised motor training with virtual reality gaming<br>Control group: supervised motor training on gaming device<br>Experimental group: MI homework 4W, 7/W, 10min + supervised training 3/W, 25min and 6W, 1/W, 10min of MI homework + supervised training 1/W, 25min<br>Control group: 4W, 3/W, 25min and 6W, 1/W, 25 min | 2 at W-1 and 1 week after intervention<br>Primary outcome: Off medication MDS-UPDRS-MS<br>3 at W0, W4 and W10<br>Secondary outcome: On medication MDS-UPDRS-MS<br>MDS-UPDRS-motor aspects of daily living<br>MDS-UPDRS-non motor aspects of daily living<br>MDS-UPDRS-summer score<br>PDQ-39 | 6 |
| Santiago et al. 2015 | RCT | Experimental group: 10 (NS), 61.30 (9.95) y, UPDRS-III 27.60 (10.04), H&Y 2.75 (range: 2-3), pharmacological treatment<br>Control group: 10 (NS), 61.40 (9.05) y, UPDRS-III 20.90 (14.85), H&Y 2.25 (range:2-3), pharmacological treatment | Modified H&Y: stage 2 to 3<br>Taking antiparkinsonian medication<br>Walking independently without any orthosis or gait-assistive device for at least 10 meters<br>Not have undergone stereotaxic surgery | NI | Experimental group: 1 session of MI + physiotherapy gait protocol<br>Control group: physiotherapy gait protocol | 4 at baseline, 10 minutes, 1 day, 7 days after training<br>Primary outcomes: stride length, total stance time<br>Secondary outcomes: hip ROM, velocity, TUG | 8 |
| Fayez et Elwishy 2013 | RCT | Experimental group: 13 (NS), 72 (3.5) y, UPDRS NS, H&Y 2.2 (0.3), pharmacological treatment<br>Control group: 13 (NS), 71 (4.2) y, UPDRS NS, H&Y 2.3 (0.3), pharmacological treatment | H&Y scale: stage 1.5 to 3<br>MMSE ≥ 26<br>Stable pharmacological treatment | Neuromuscular problems that affected their motor performance<br>Vestibular dysfunction<br>H&Y scale ≥ stage 4 | Experimental group: physiotherapy + MI of gait<br>Control group: physiotherapy + watching documentaries. Both groups: 4W, 3/W<br>Physiotherapy: callisthenic exercises (15-20min), practice of specific functions for lower and upper limb (15-20 min) and relaxation exercises<br>MI of gait and documentaries: 25-30 minutes | 2 W0 and W4<br>Step length, Walking velocity<br>Excursions in sagittal plane of the ankle, knee, hip joints<br>FGA | 7 |
| Braun et al. 2011 | RCT | Experimental group: 25 (8♀), 70 (8) y, UPDRS NI, H&Y (range: 1-4), NI<br>Control group: 22 (7♀), 69 (8) y, UPDRS NI, H&Y (range: 1-4), NI | Clinically diagnosed adults with Parkinson's disease<br>Being able to engage in mental practice (clinical judgment of the treating therapist, support from family, MMSE score) | Other conditions such as stroke<br>Rheumatic diseases<br>Dementia prior to the onset of Parkinson's disease and sufficient to cause persistent premorbid disability | Experimental group: physiotherapy + MI of locomotor tasks adapted for each participant<br>Control group: physiotherapy + relaxation (sham intervention)<br>For both groups: 6W<br>Physiotherapy: 1h or 2 times 30 min<br>MI: 20 min or 2 times 10 min | 3 at W0, W6 and W12<br>VAS for gait improvement (0 'poor' and 10 'excellent')<br>TUG<br>10MWT | 8 |
| Tamir et al. 2007 | RCT | Experimental group: 12 (4♀), 67.4 (9.7) y, UPDRS NI, H&Y 2.29 (0.4), pharmacological treatment<br>Control group: 11 (4♀), 67.4 (9.1) y, UPDRS NI, H&Y 2.31 (0.4), pharmacological treatment | Community-dwelling individuals with PD<br>H&Y scale: stages 1.5 to 3<br>MMSE ≥ 26 points | Presence of neuromuscular or skeletal comorbidities that affected their motor performance<br>H&Y scale: stage 4<br>Ailments that prevented from making moderate physical efforts | Experimental group: physiotherapy + MI practice<br>Control group: physiotherapy only<br>For both groups: 12W, 2/W, 1h<br>Physiotherapy: callisthenic exercises (15-20 min), practice of specific functions for lower limb and upper limb (15-20 min), relaxation exercises<br>MI practice: integrated in physiotherapy, either preceded the motor task or followed it<br>Tested in on-state | 2 at 1 day before and at the end of the intervention<br>TUG<br>Standing up and lying down<br>Turning in place 360 deg<br>Tandem stance<br>Functional reach test<br>Shoulder tug<br>UPDRS<br>Clock drawing<br>Stroop test | 6 |

**Abbreviations:** ♀, woman/women; ♂, man/men; 10MWT, 10-Meter Walking Test; ABC, Activities Balance Confidence; AOT, Action observation therapy; DGI, Dynamix Gait Index; FES-I, Falls Efficacy Scale International; FGA, Functional Gait Assessment; H&Y, Hoehn and Yahr; HS, healthy subjects; KVIQ-20, Kinesthetic and Visual Imagery Questionnaire-20; MDS-UPDRS(-MS), Movement Disorder Society-Unified Parkinson Disease Rating Scale-(Motor Scale); MI, motor imagery; MiniBESTest, Mini Balance Evaluation Systems Test; MMSE, Mini Mental State Examination; MoCA, Montreal Cognitive Assessment; NFOG-Q, New Freezing of Gait Questionnaire; NI, non-informed data; PD, Parkinson disease; PDQ-39, Parkinson's Disease Questionnaire-39 items; SD, standard deviation; TUG, Timed Up and Go; W, week(s) ;Y, years; ROM, range of motion; VAS, visual analogue scale;

Most studies had as inclusion criteria a H&Y score ≤3 (23–25,31,32,34–38) except for Sarasso et al. (33) who included patients with a H&Y score ≤4. One study (31) did not report eligibility criteria related to an H&Y score and one study (31) excluded patients with H&Y score >3. For the exclusion criteria, in most studies, patients with neuromuscular, psychiatric, or neurological pathologies other than PD were excluded.

#### 3.2.2 Protocols

Regarding the 12 RCTs, the mean protocol duration was 7 weeks, ranging from a single session to 12 weeks with a mean number of sessions per weeks of 3 (range: 1–7). Duration of the interventions was specified for 7 studies, with a mean duration of 55 minutes for experimental group (range: 35–80) and 52 minutes for control group (range: 25–80). All studies performed a pre-intervention and post-intervention assessments and 3 studies (28,30,32) included a follow-up intervention ranging from one week to 8 weeks after the end of the protocol. Concerning the types of exercises, eight studies (22–24,28–30,32,33) used a MI protocol of gait and balance exercises or gait exercises only. One study (34) included a protocol of MI of a single step. Two studies (32,36) used a guided neurofeedback protocol with MI.

#### 3.2.3 Outcomes

In terms of motor symptoms, two studies (32,36) used Movement Disorder Society (MDS) Unified Parkinson’s Disease Rating Scale (UPDRS) as primary outcomes. They compared part III of UPDRS. Regarding the assessment of quality of life, only 4 studies (23, 24, 31,36) assessed this parameter using the Parkinson’s Disease Questionnaire-39 (PDQ-39). Walking and balance abilities were assessed including walking speed, step length, Timed Up and Go (TUG), Dynamic Gait Index (DGI), Functional Gait Assessment (FGA), 10-Meter Walk Test (10MWT), 2-minute endurance walking test, sit-to-stand, balance test (23–25, 31, 33–37). TUG test was used in 6 studies as outcomes (25, 31,33–35, 37). Six studies focused on balance (23–25, 33–35). Lower limb range of motion (ROM) was also assessed in two studies, one (35) focusing on hip and other (36) evaluated hip, knee and ankle. No specific upper limb or speech outcomes have been assessed.

#### 3.2.4 Results of RCT

Intergroup significant differences range were very large among the 12 studies, 10 showed a significant difference between groups at post intervention (Table 2).

**Table 2.**
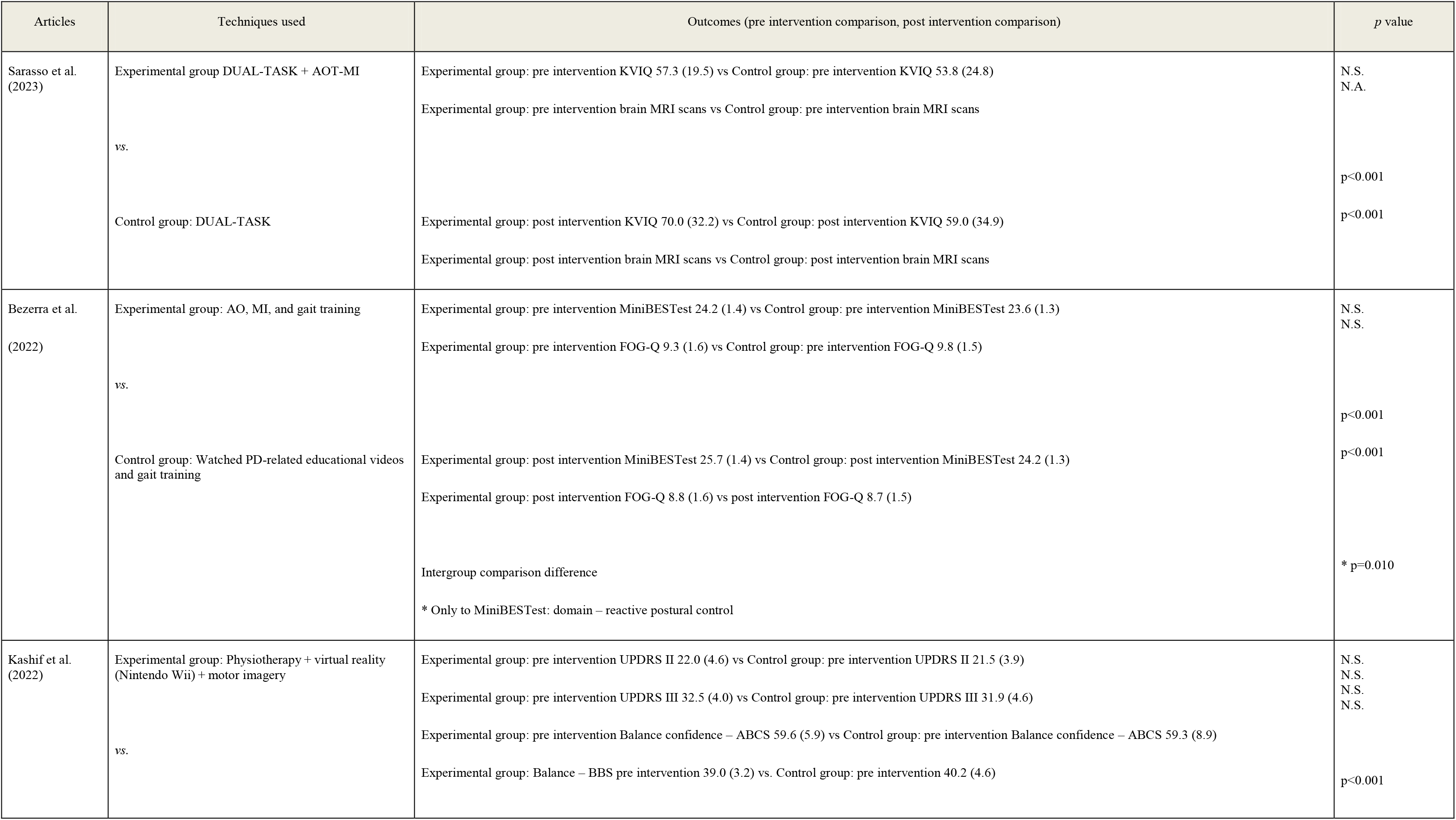
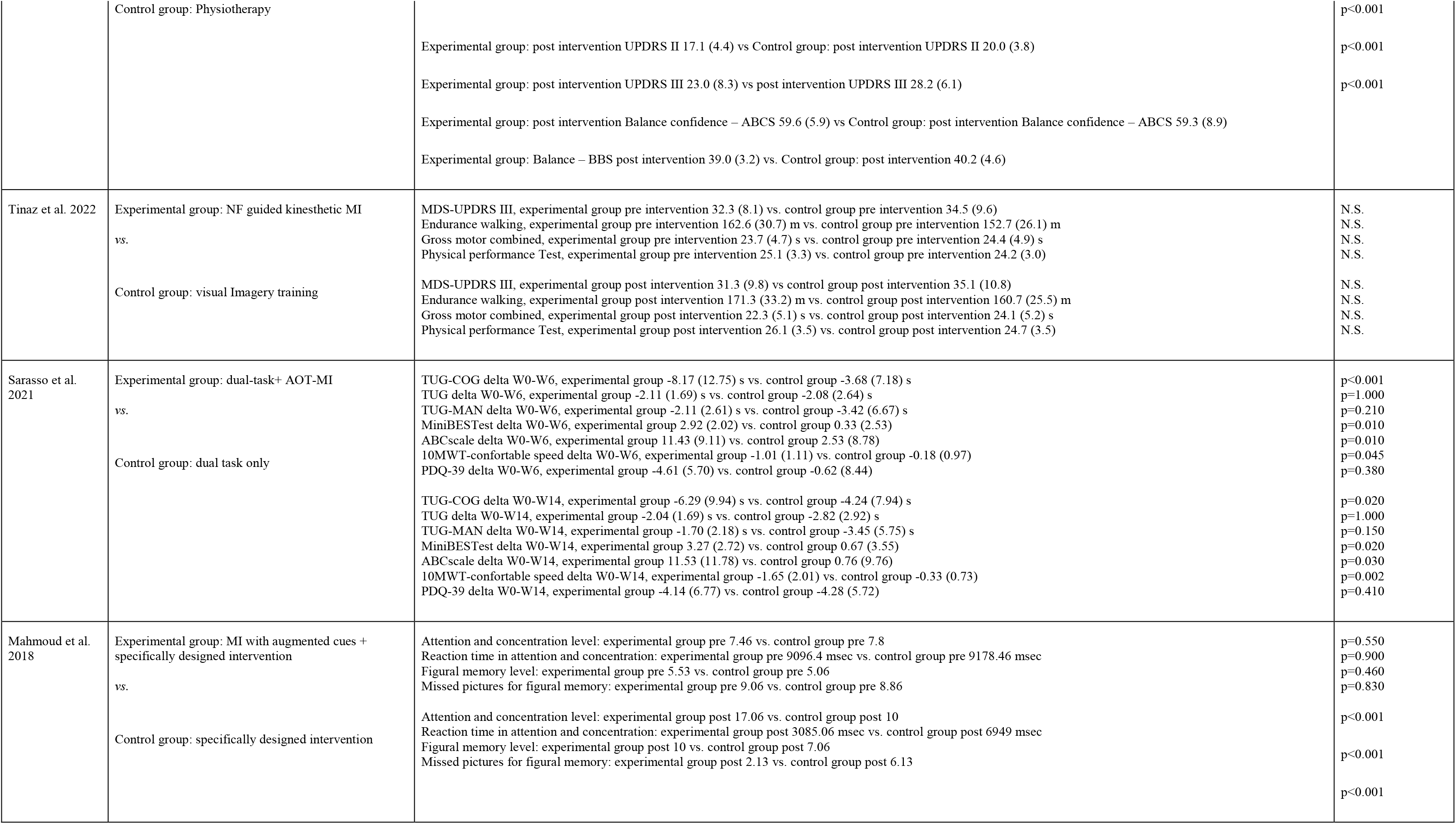
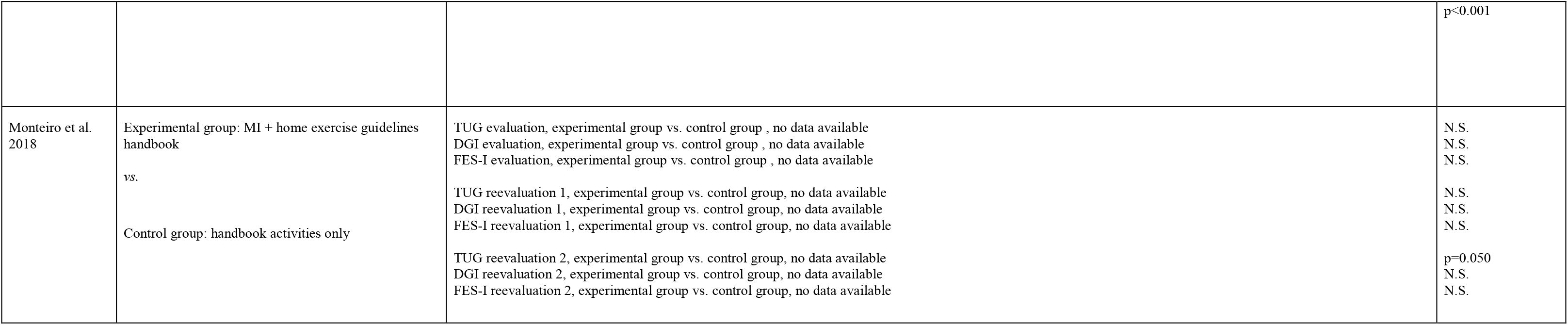
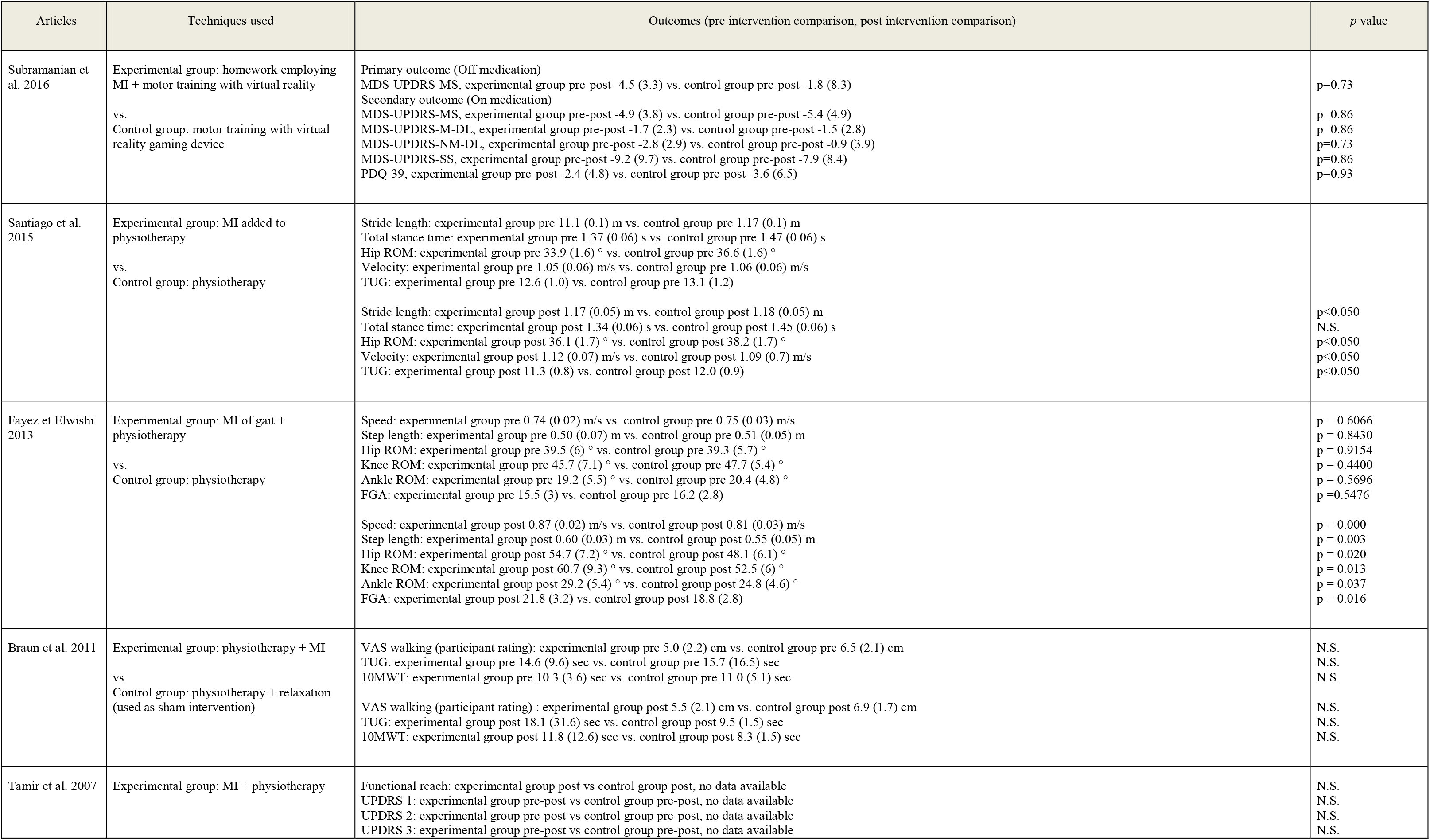
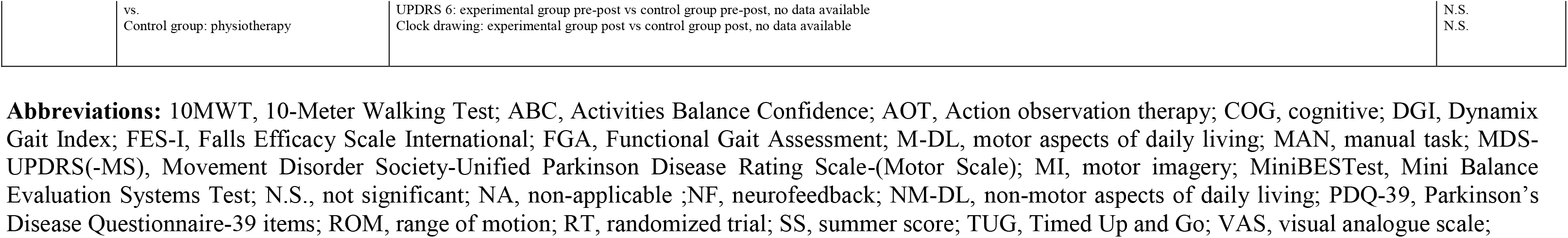
Results of randomised controlled trials.

Regarding the studies with a **gait and balance MI exercises**, Sarasso et al. (33) found a significant improvement in TUG with a cognitive task (primary outcome) compared to control group. An improvement of 122% (p<0.001) was found in the week 6, and 48.3% (p=0.02) in the week 14. Santiago et al. (35) also found an improvement in TUG for the experimental group (5.8%; p<0.05). Sarasso et al. (33) showed an improvement of 388.05% (p=0.020) at week 14 for the experimental group for the Mini Balance Evaluation System Test as well as an improvement of 1417.1% (p=0.03) for the Activities-specific Balance Confidence Scale. Mahmoud et al. (32) focused on concentration parameters. The level of attention and concentration was significantly improved by 70.6% (p<0.000). The reaction time of the previous test was also improved by 55% (p<0.000). Two other variables on figural memory were improved (range: 42–65%; p<0.000). Fayez and Elwishi (36) found a significant difference for hip, knee, and ankle ROM in the experimental group (range: 13.7–17.7%; p<0.013– 0.037). For the spatiotemporal parameters, Fayez and Elwishi (36) showed a significant improvement in walking speed by 7.4% (p<0.000), step length by 9.1% (p=0.002) and FGA by 16% (p<0.016) in the experimental group. Santiago et al. (35) showed a significant improvement in walking speed (2.8%; p<0.05) in the experimental group. Sarasso et al. (33) showed an improvement of 400% at week 14 for the 10MWT. Monteiro et al. (37) studied **MI on only one step execution** and they showed a significant difference for the TUG test at 14 week (difference not specified; p=0.05).

Regarding the MI exercises studied (**neurofeedback protocol, gait and balance, step**) no significant differences was found between experimental and control group for the follow outcomes: TUG, hip ROM, step length, 10MWT, MDS-UPDRS score, endurance walking, gross motor combined, physical performance test, PDQ-39, DGI, FGA, Falls Efficacy Scale International, Functional Reach Test, total stance time (25,31,33–35,37,38).

Another interesting result was found by Sarasso et al. (2023), where the MI was assessed using Kinesthetic-and-Visual-Imagery Questionnaire (KVIQ) and a MI functional MRI (fMRI) task. During fMRI, subjects were asked to watch first-person perspective videos representing gait/balance tasks and mentally simulate their execution. At baseline patients were compared with 23 healthy controls. They showed that observation and MI training (AOT-MI) in PD patients promoting the functional plasticity of brain areas involved in MI processes and gait/balance control (22). There are no results for upper limb and speech as no specific outcomes were assessed.

### 3.3 Non-RCTs and descriptive studies: assessment of MI and main results

The results of the following studies should be interpreted with caution as we focused only on their main results. As far as possible, we have organised the results according to this logic: firstly, the difference between patients with PD and healthy subjects (HS) in terms of MI (PD/HS-MI); secondly, the difference between patients with PD and HS in terms of ME (PD/HS-ME); and finally, the difference between ME and MI (MI/ME) for a same group of patients. The characteristics of the descriptive and non-RCTs studies are in Table 3 and the main results in Table 4.

**Table 3.**
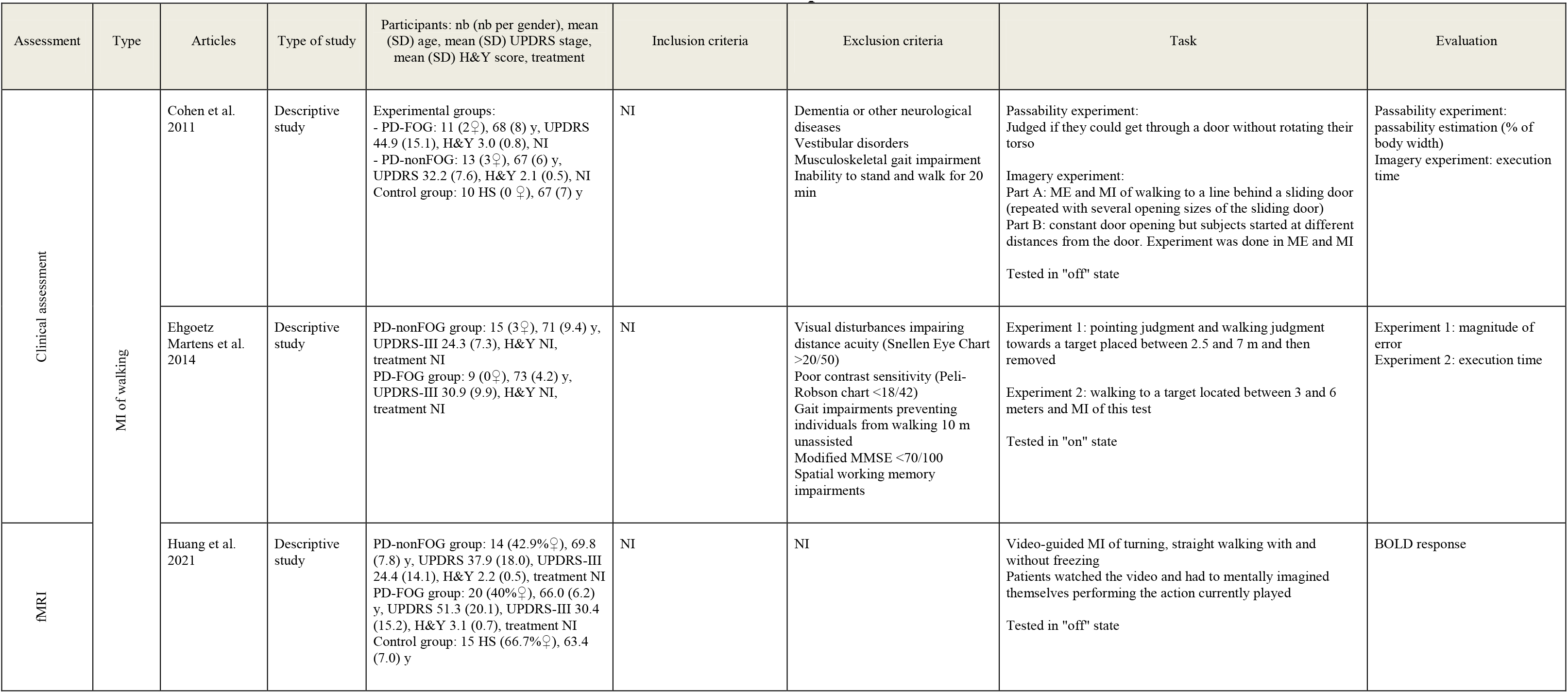
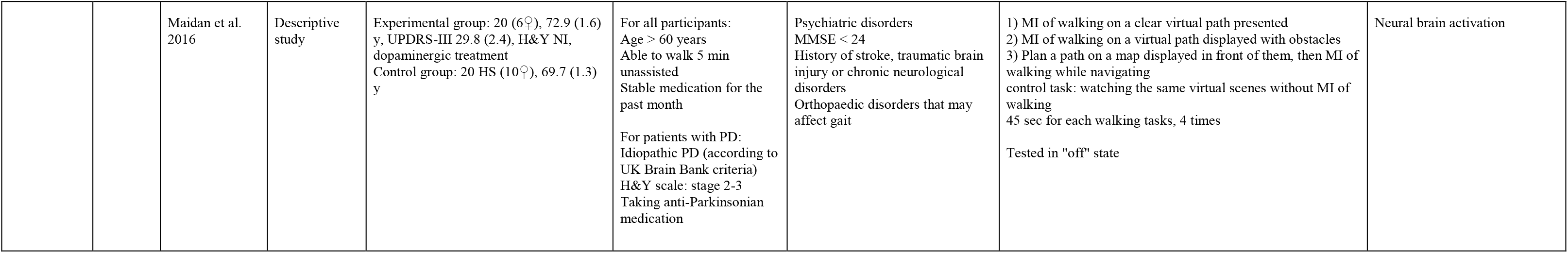
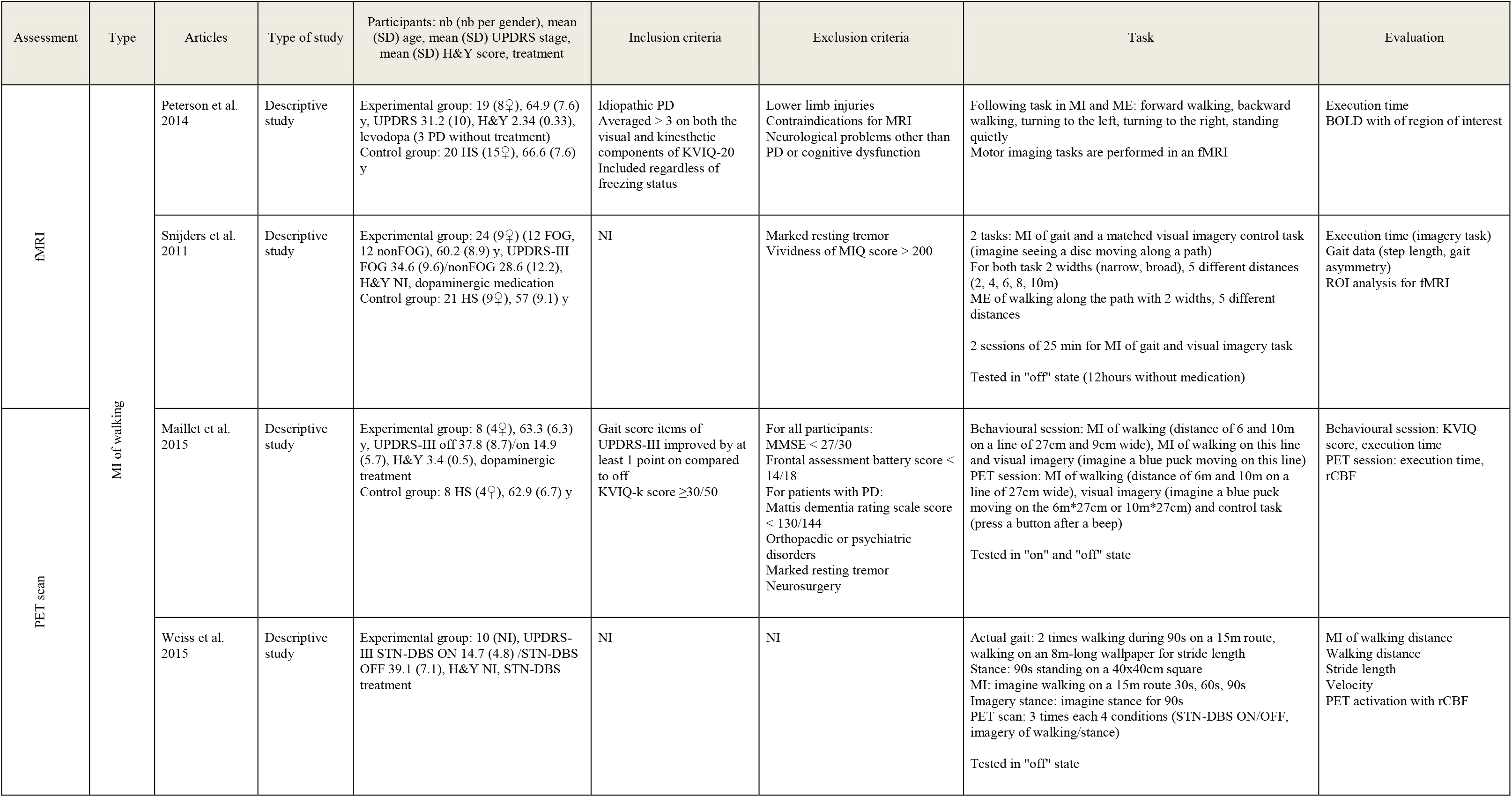
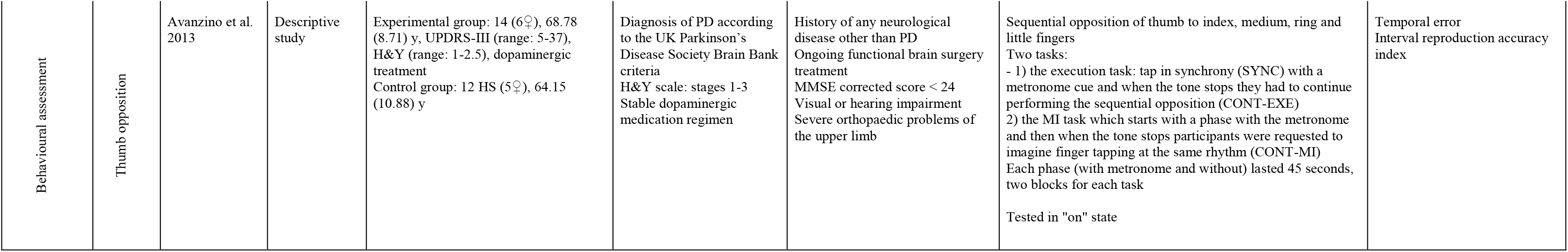
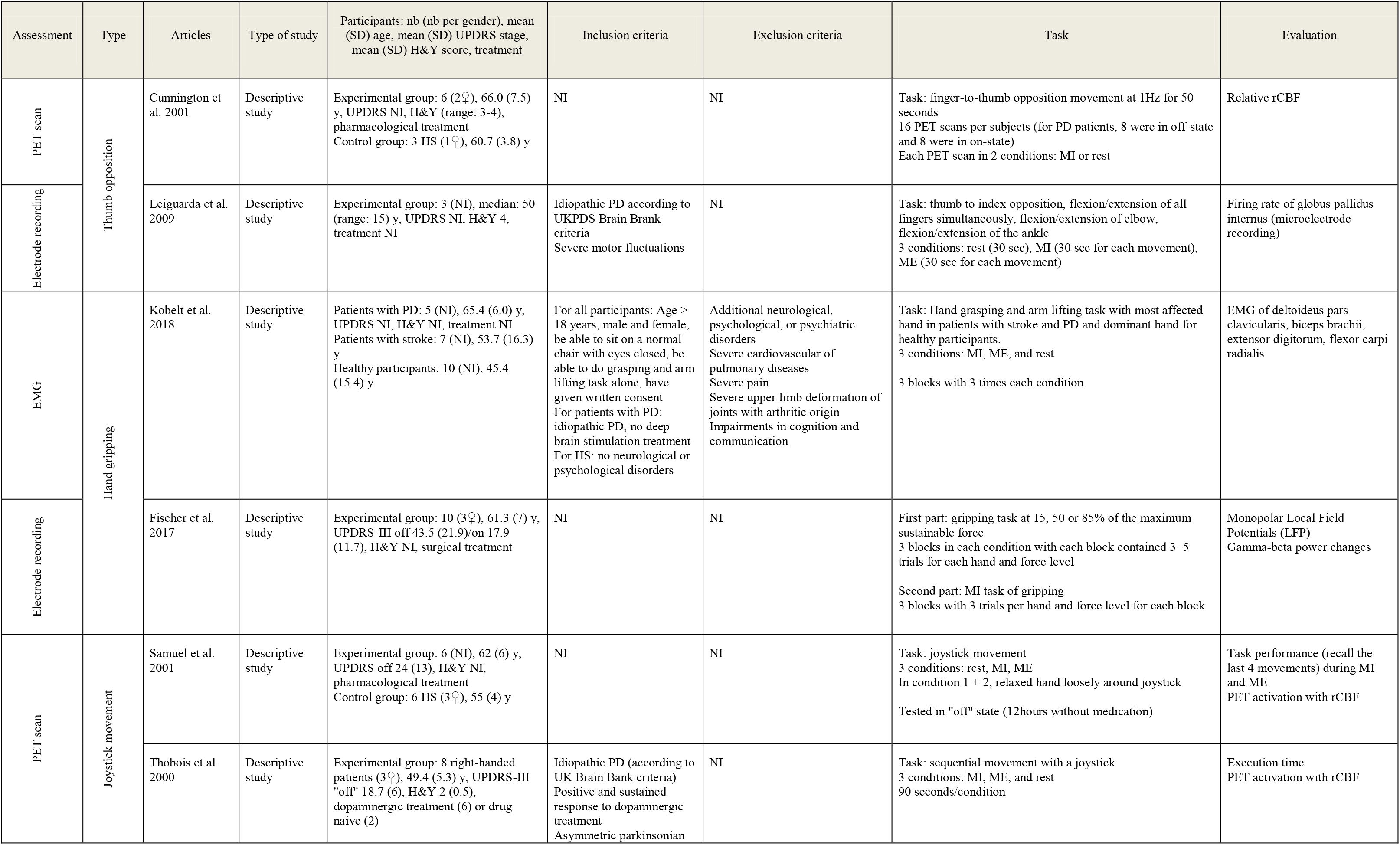
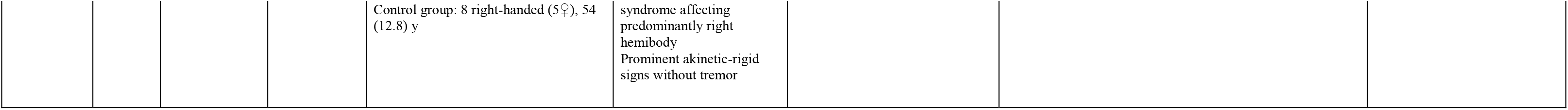
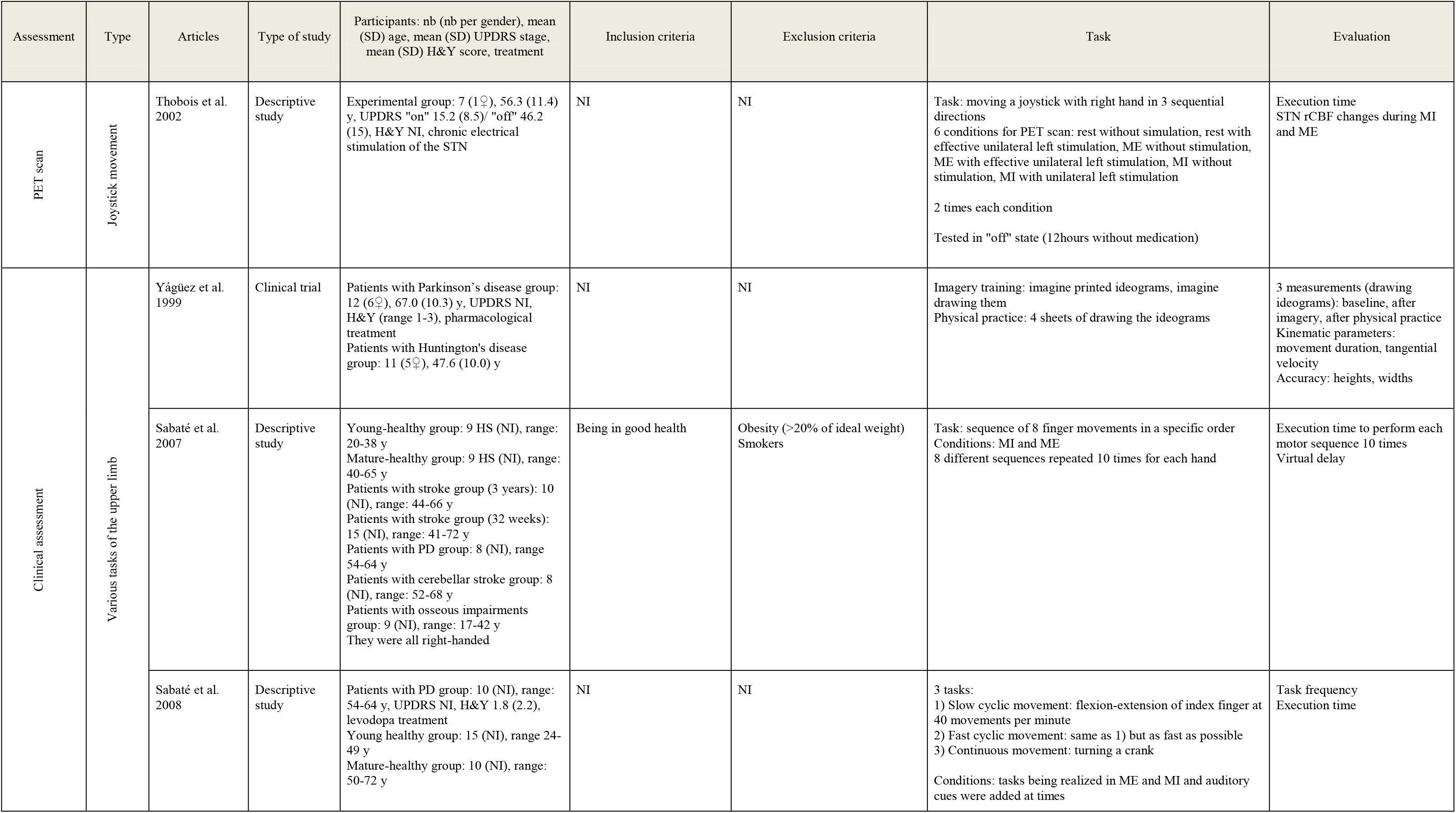
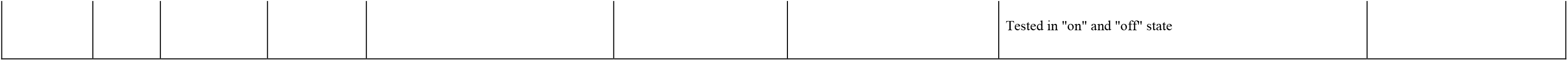
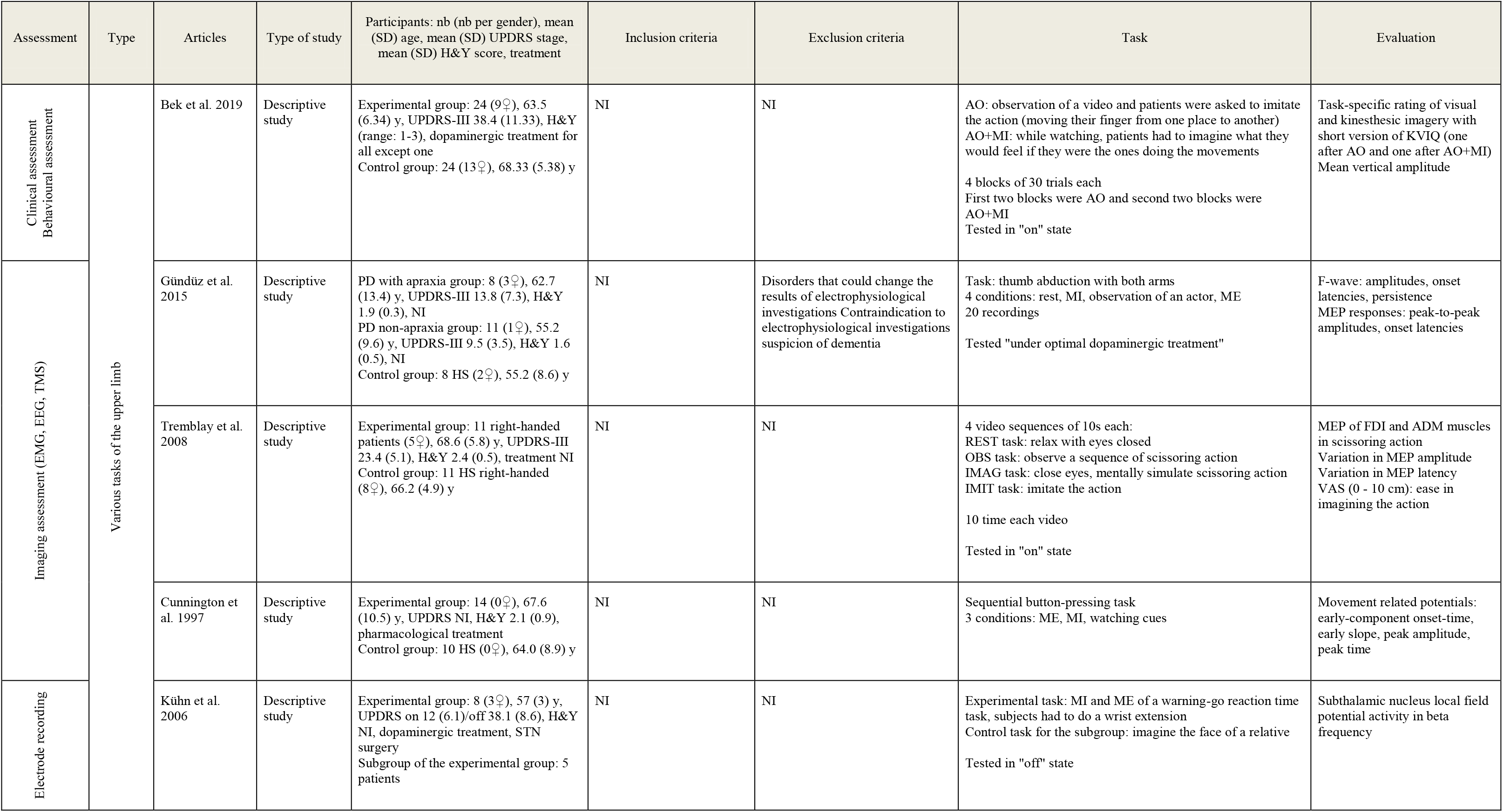
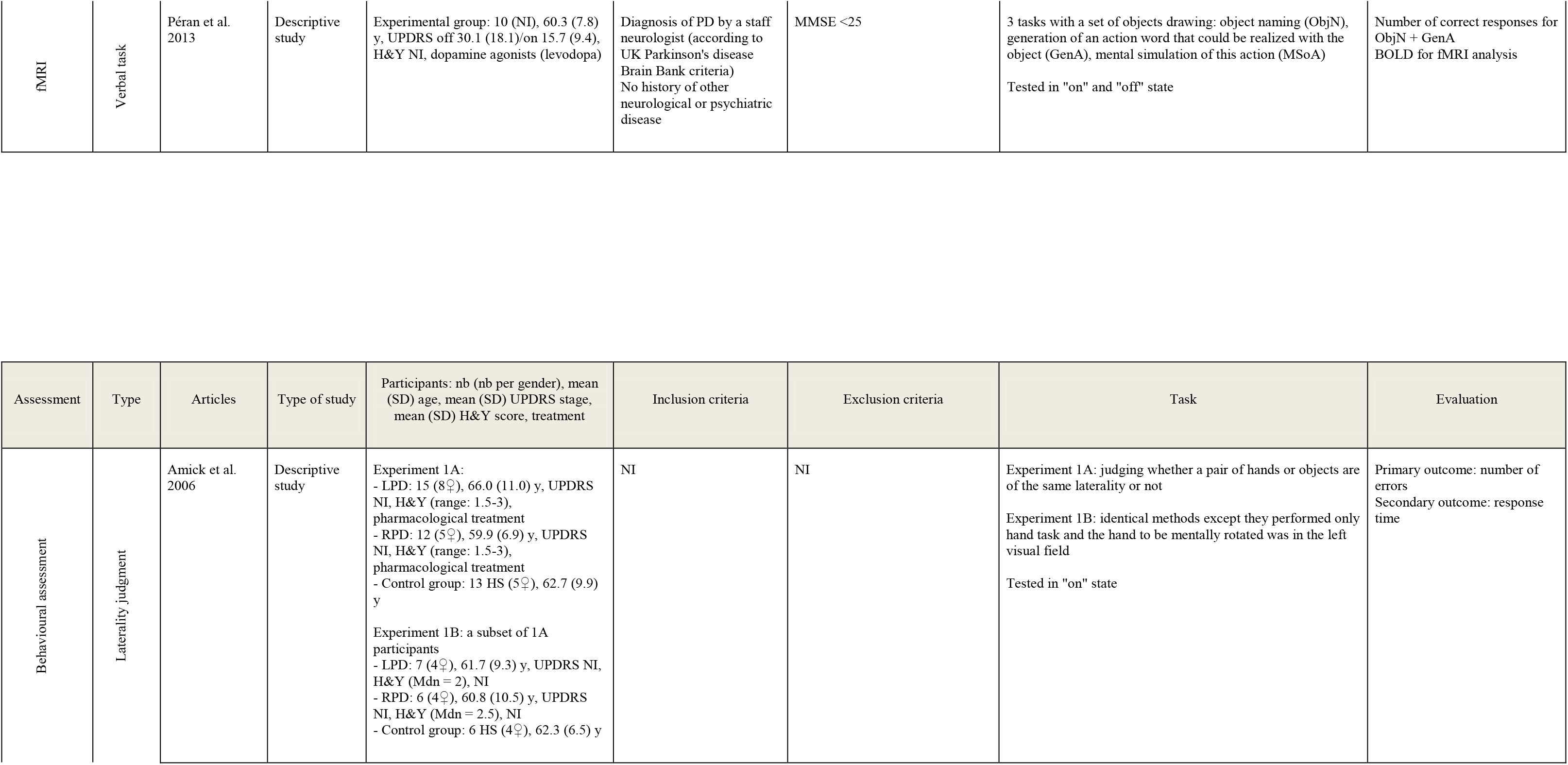
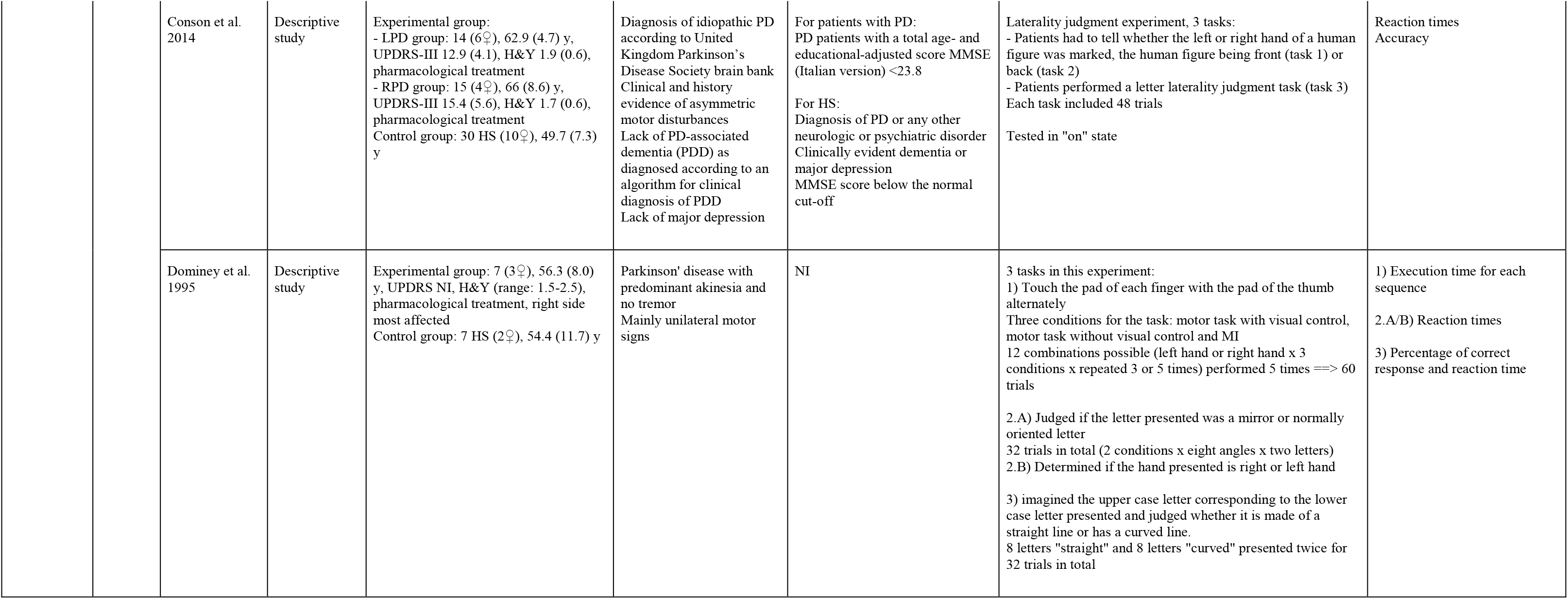
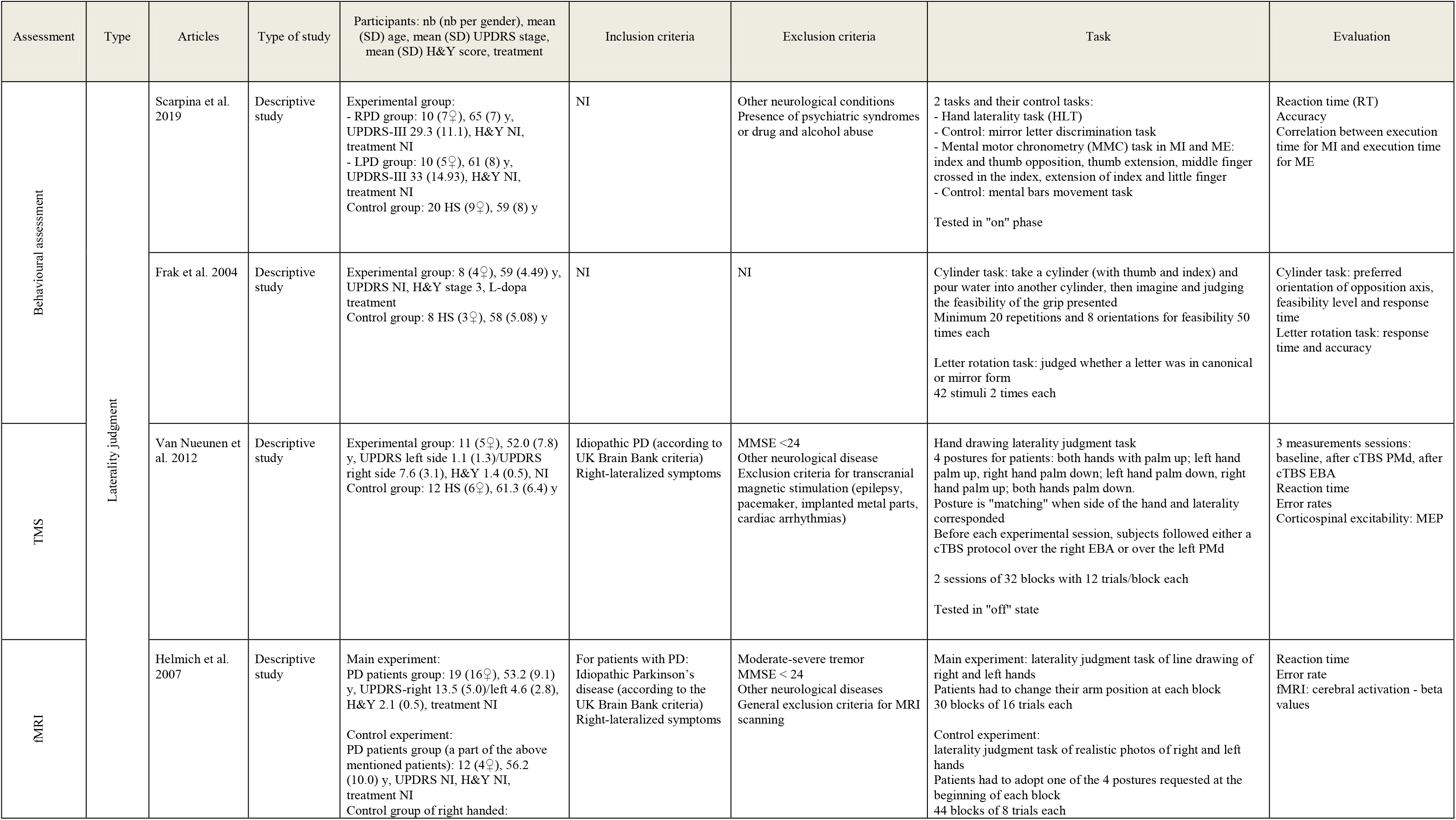
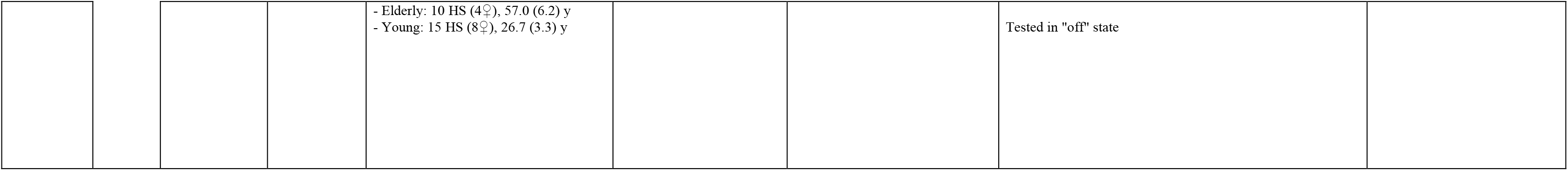
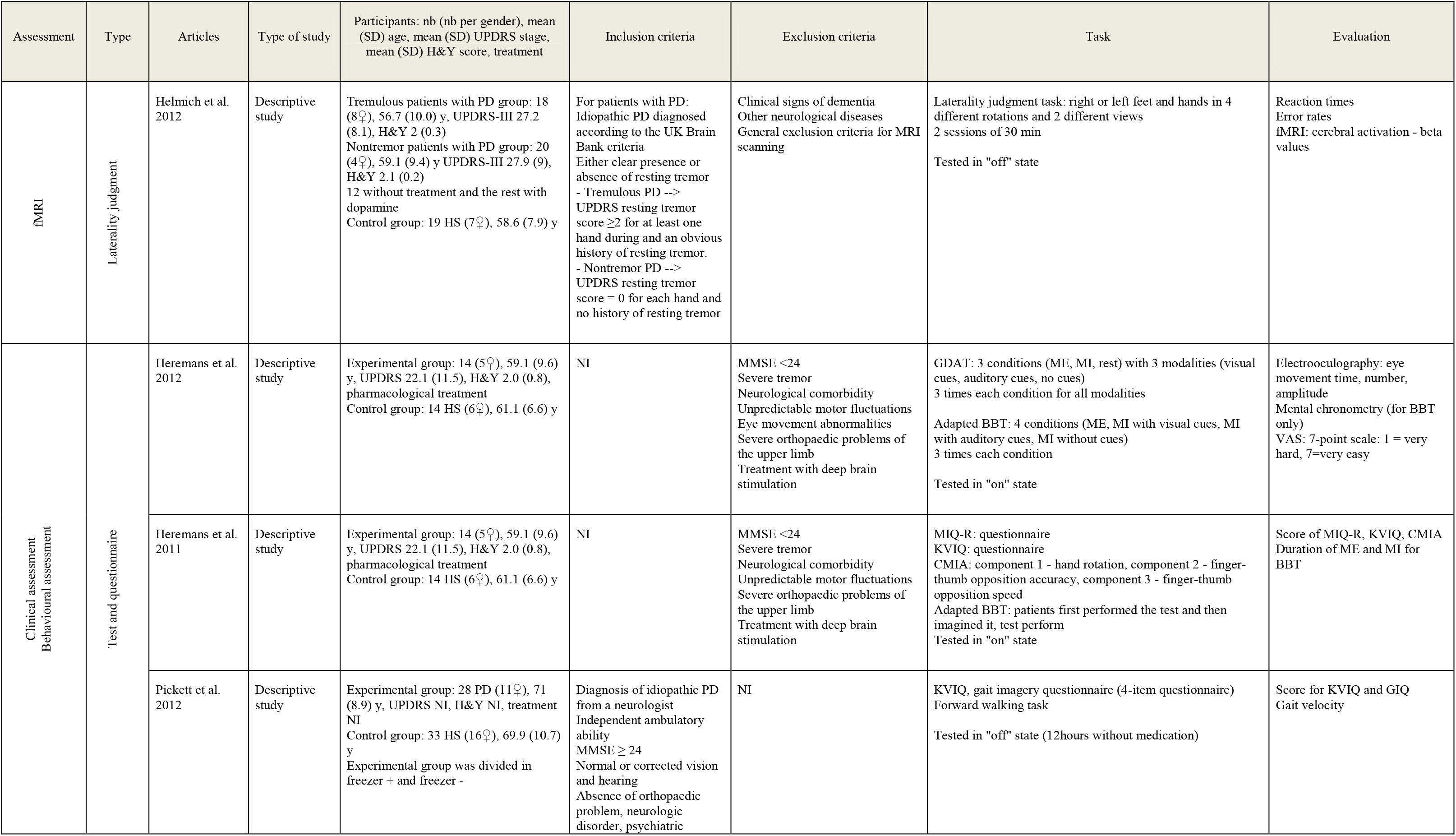
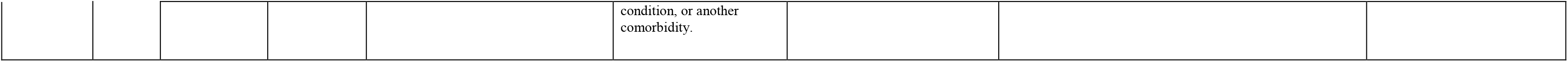
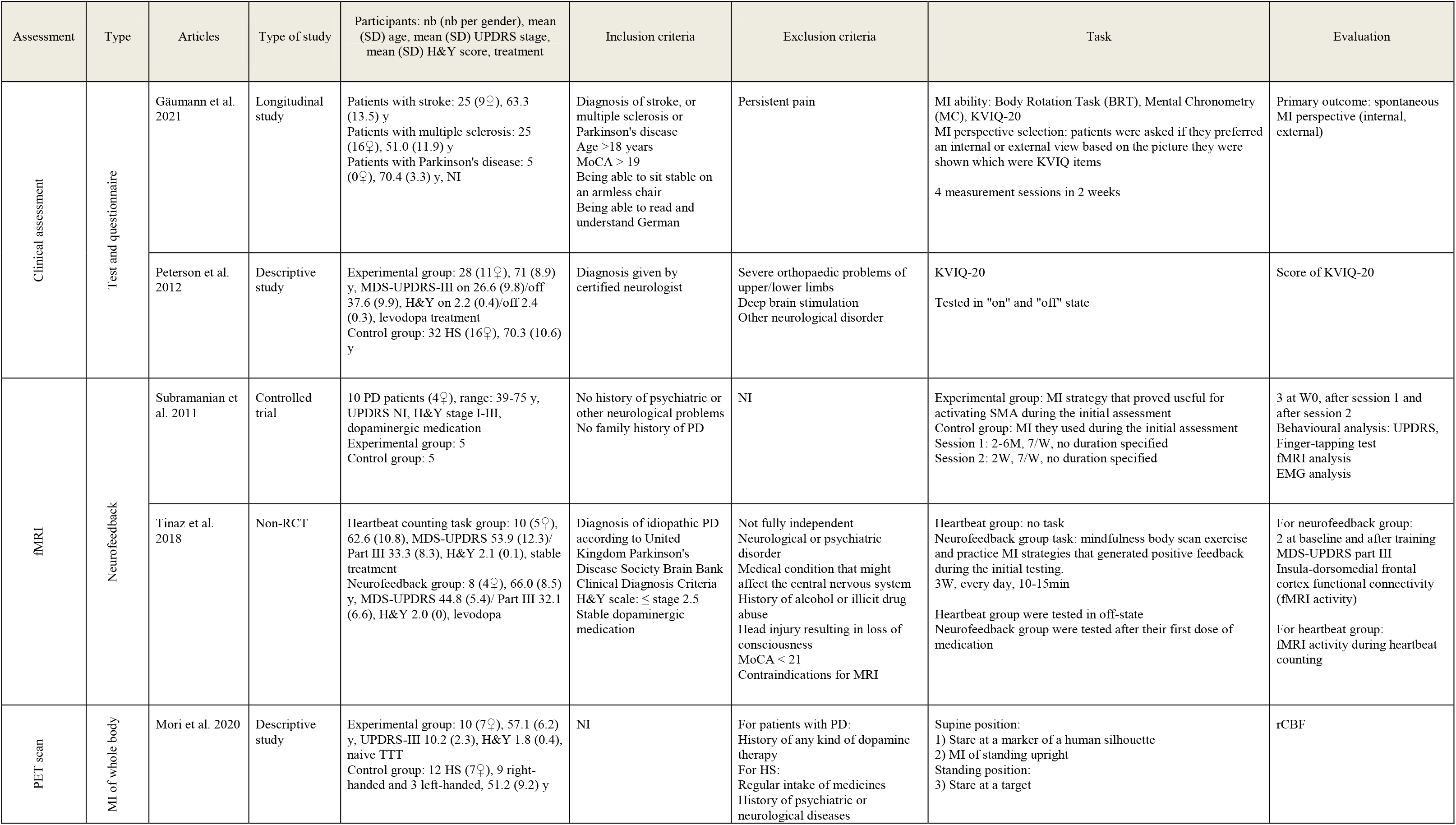
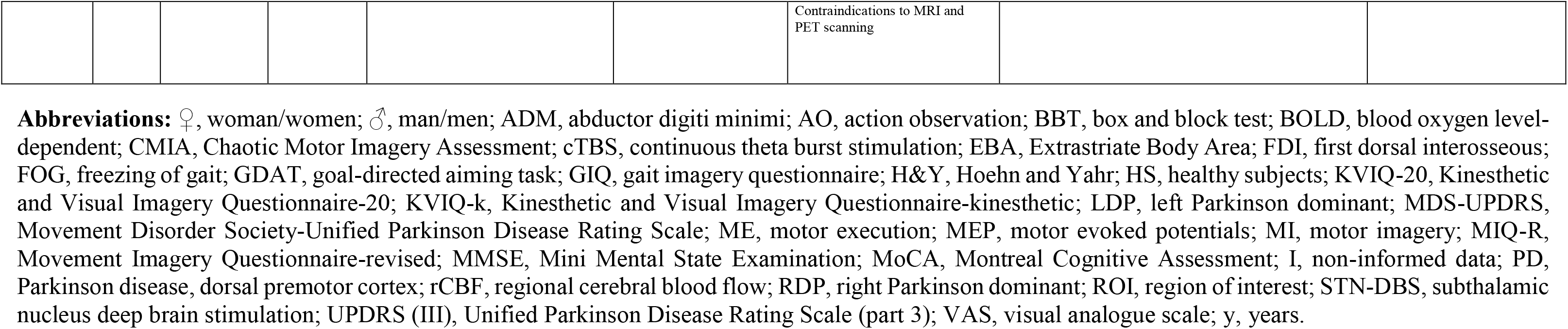
Characteristics of non-randomised controlled trials and descriptive studies.

**Table 4.**
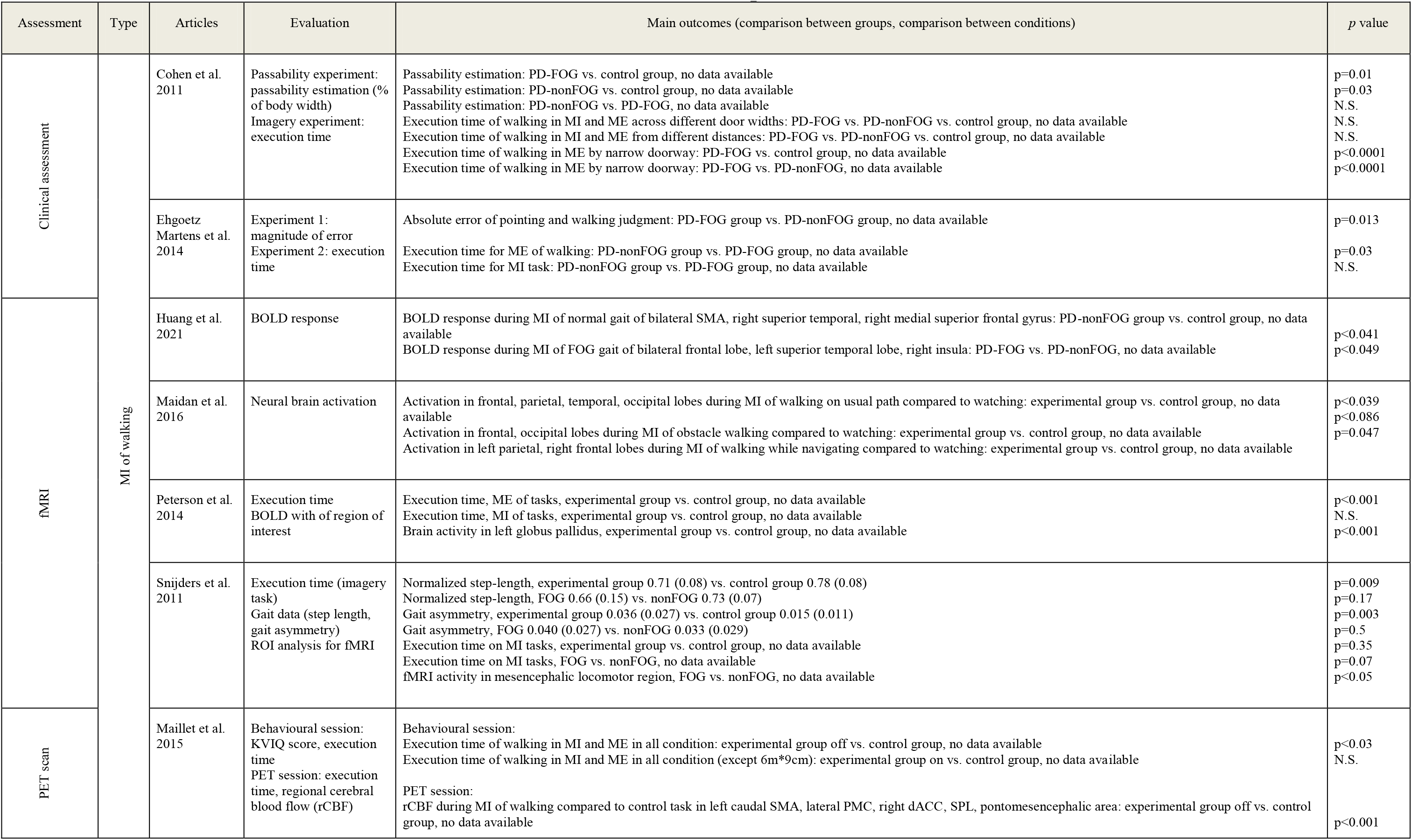
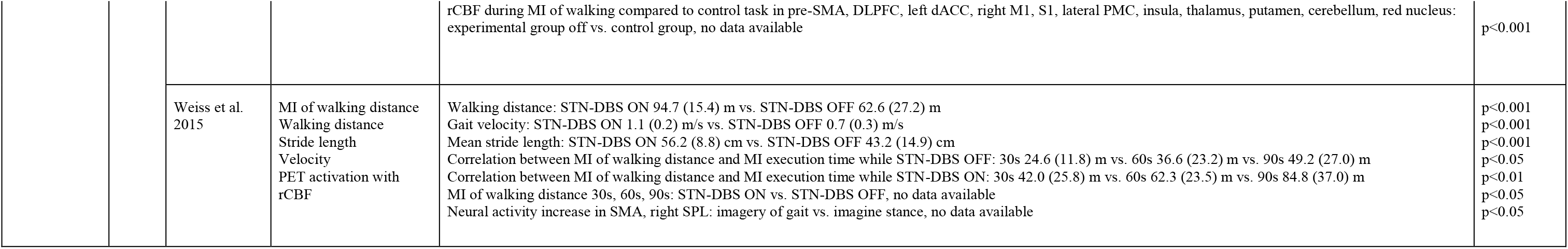
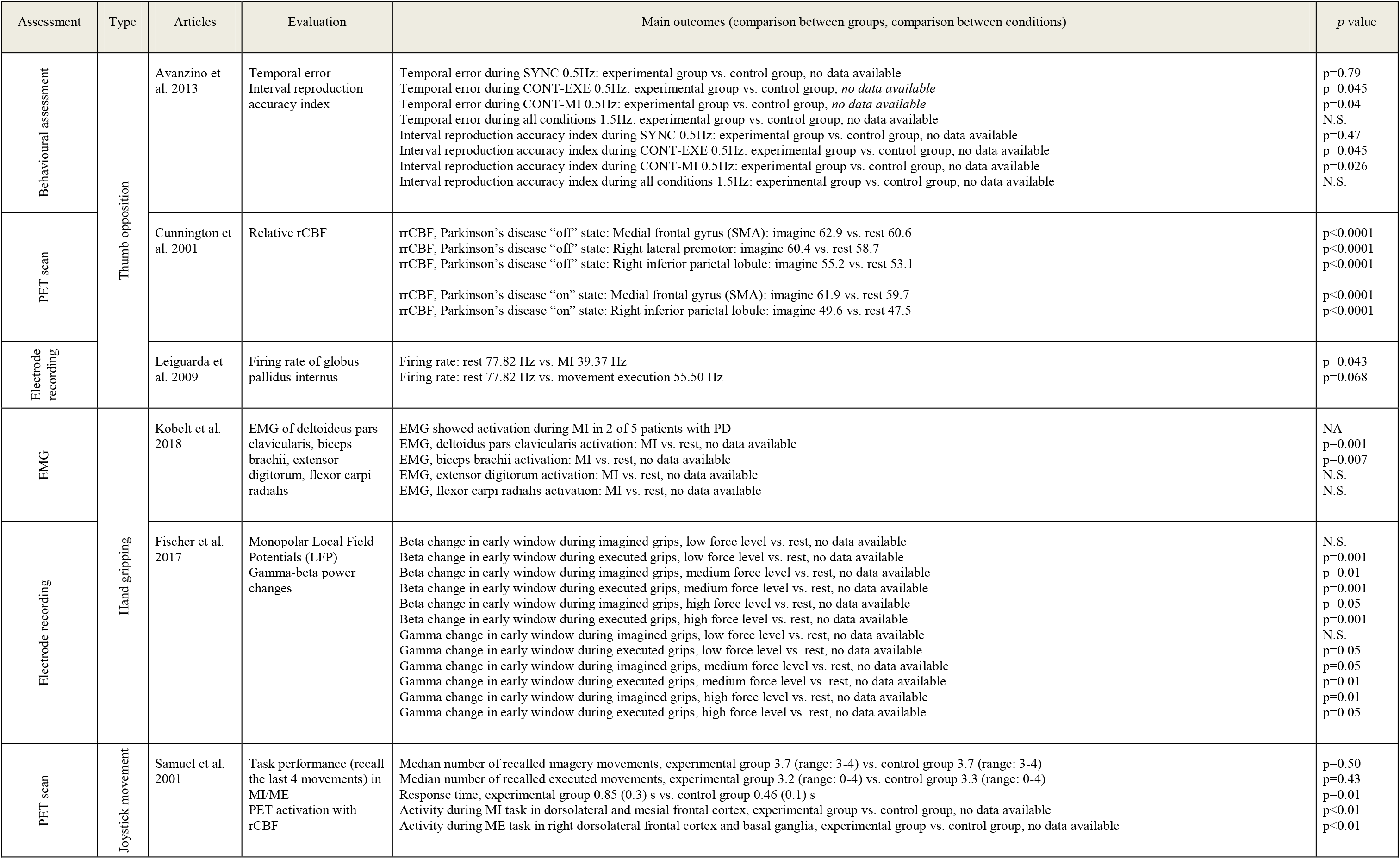
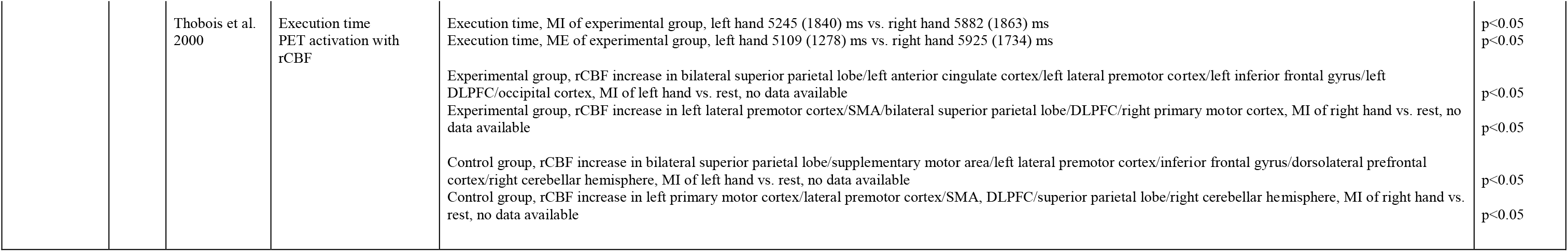
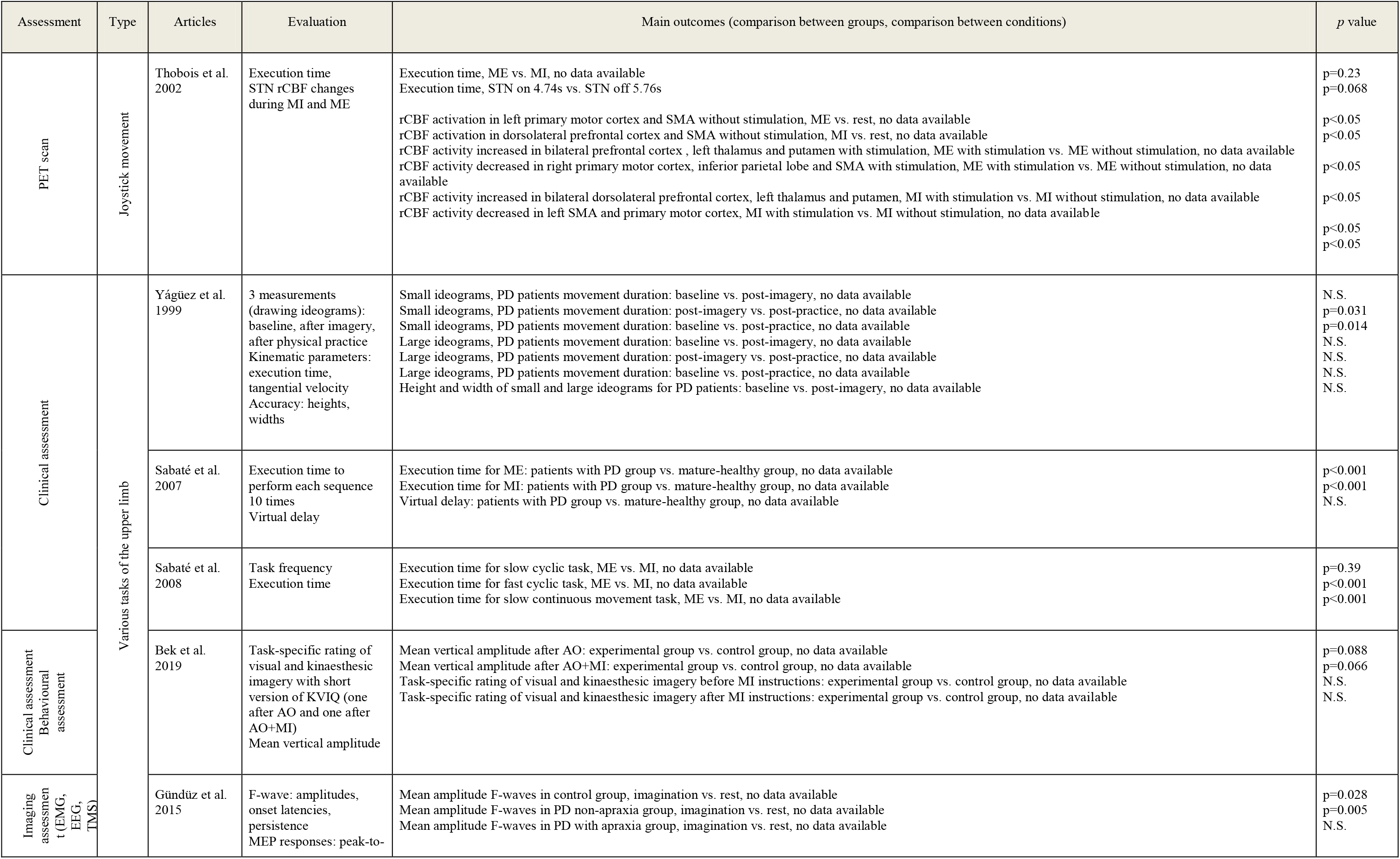
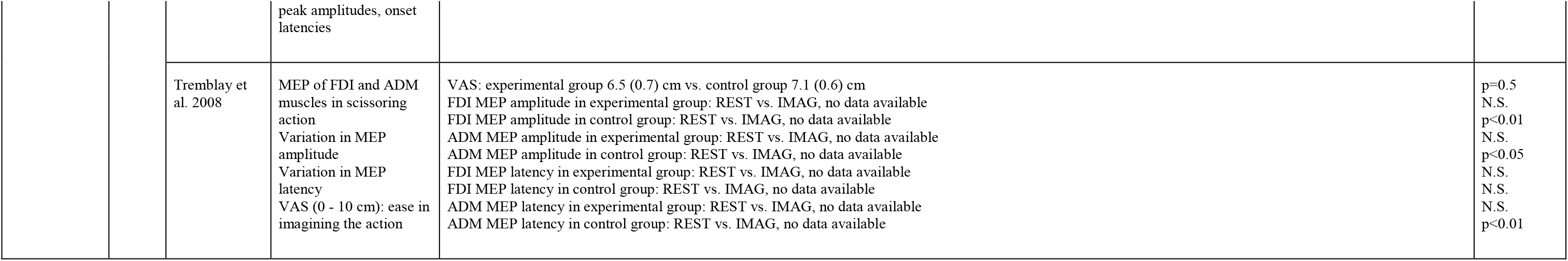
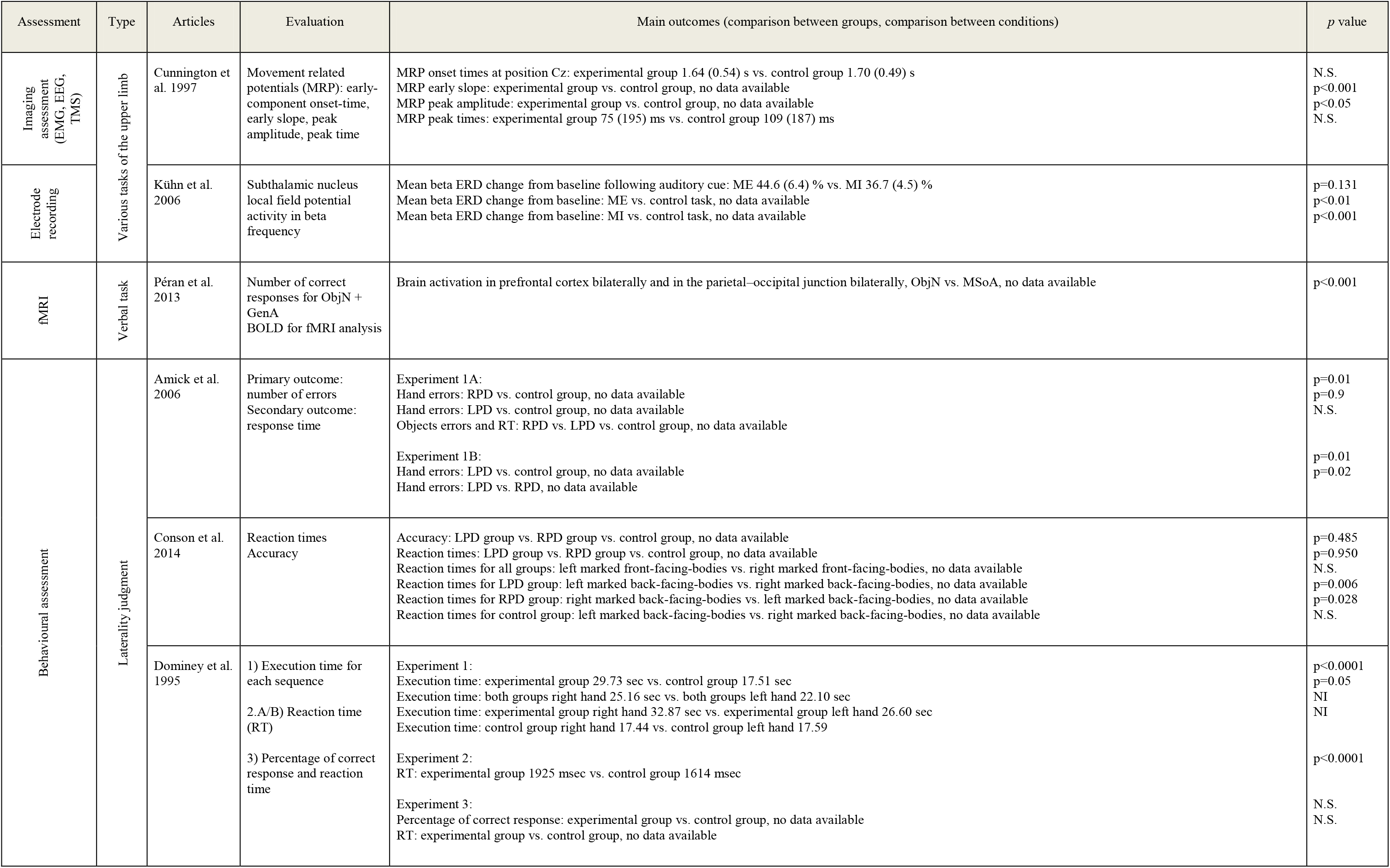
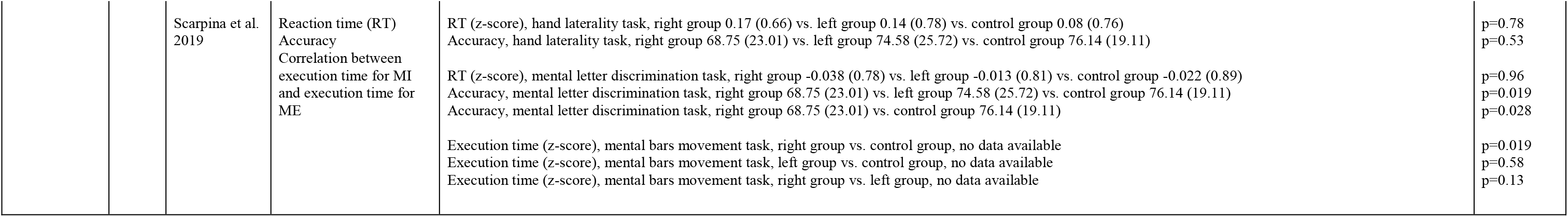
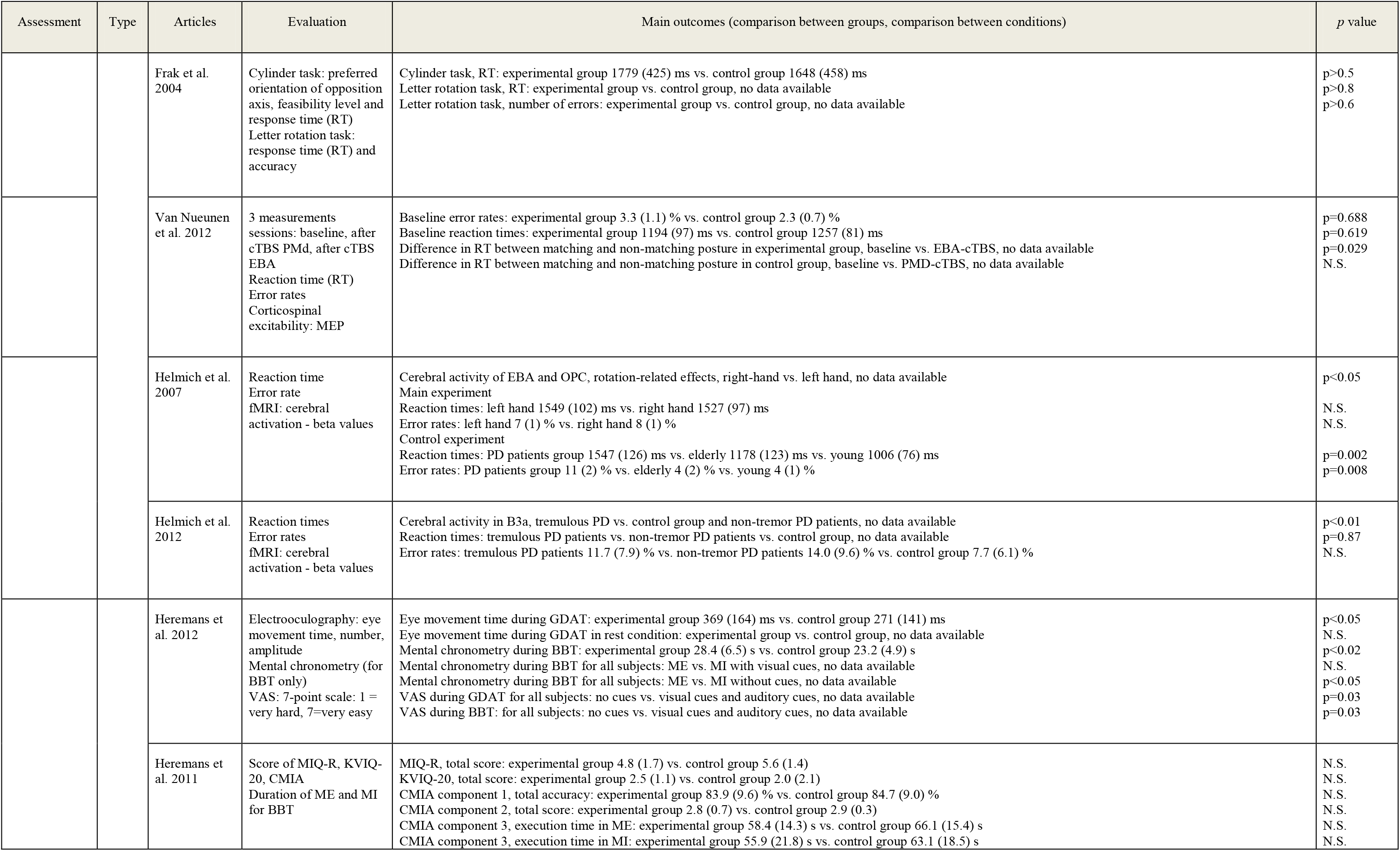
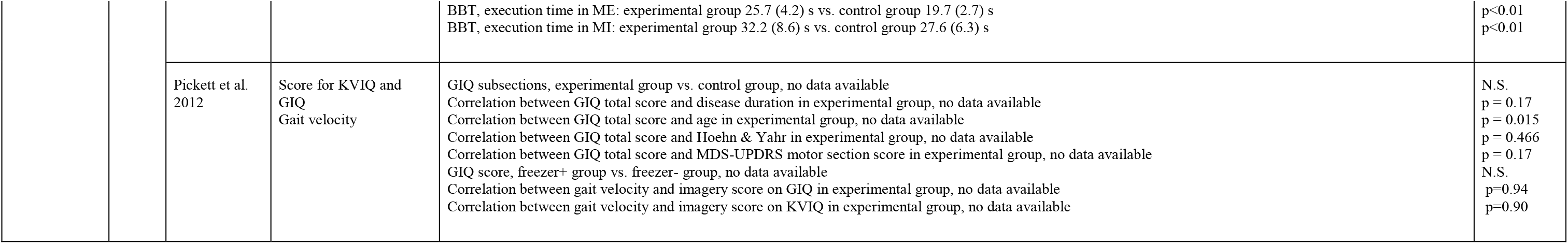
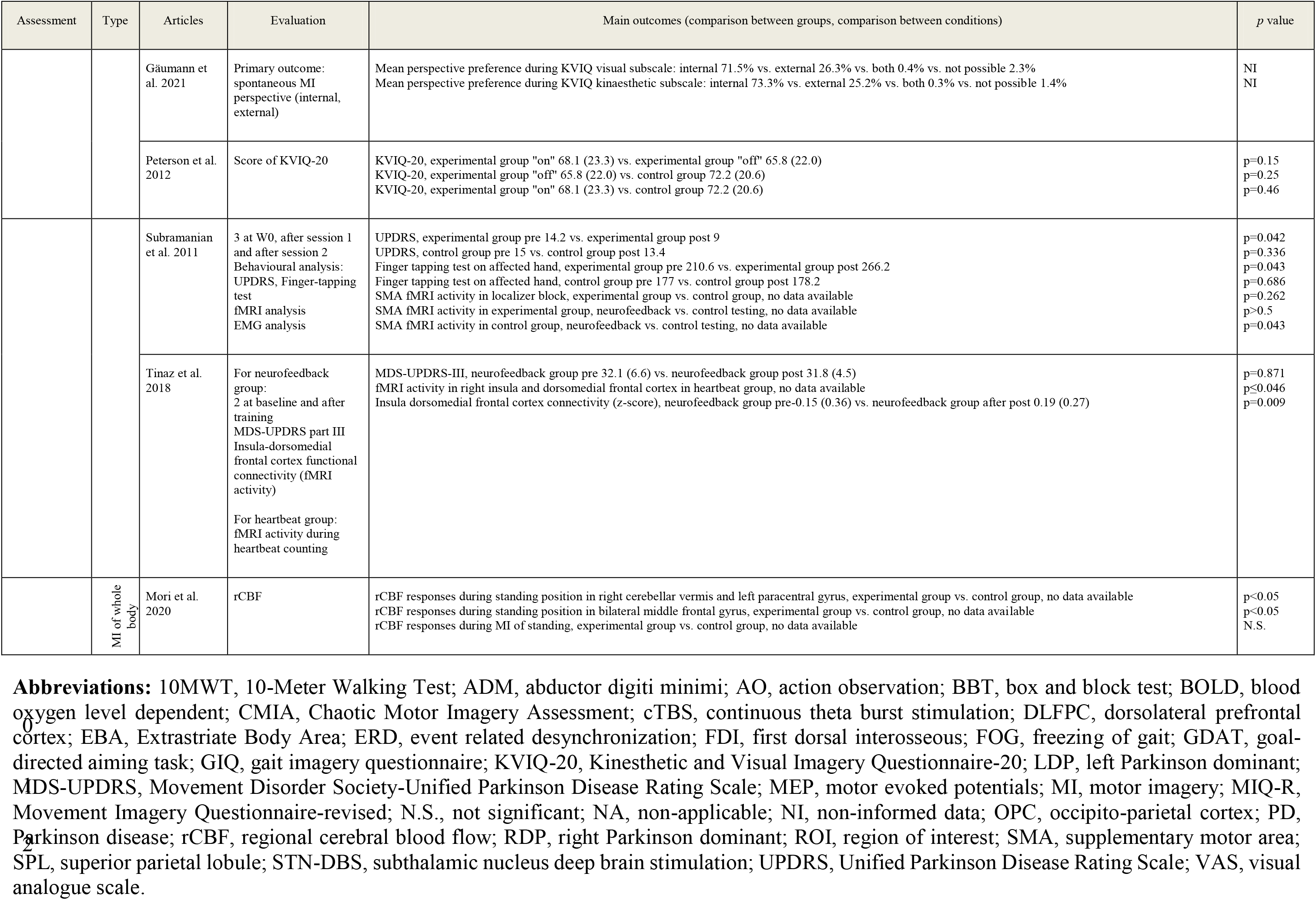
Main results of non-randomised controlled trials and descriptive studies.

#### 3.3.1 Participants characteristics

Most of these studies (39–41), patients with PD were compared with HS of the same age. The mean (SD) number of participants per study was 30 (±18) and participants had a mean age of 61 (±8) years old. Groups were composed on average of 35.5% of women and 64.5% of men. For patients with PD, the main inclusion criteria were a diagnosis of idiopathic PD (10 studies specified that the diagnosis was made with the UK brain bank criteria) and the H&Y score. Twenty-one out of 41 studies did not mention inclusion criteria. Four studies included patients with other neurological conditions such as stroke, multiple sclerosis, Huntington’s disease (39–42).

Regarding the inclusion criteria for MI, the Kinesthetic and Visual Imagery Questionnaire (KVIQ) which evaluates the ability of subjects to imagine from a first-person perspective by assessing the clarity of the image (visual: V subscale) and the intensity of the sensations (kinesthetic: K subscale) was used.

#### 3.3.2 Protocols

We have grouped the studies according to whether they concerned the lower limb, the upper limb or language-related MI exercises. Subgroups were made within these categories.

Eight studies concerned the lower limb through MI of walking. Among these studies the protocols were heterogeneous. Five studies tested MI walking in a straight line with different distances ranging from 2 to 15 m; 2 studies tested MI walking in a straight line, turning, turning back; and 1 study tested walking on an obstacle path.

Upper limb was involved in 16 studies. Three studies tested a thumb opposition task; 2 studies tested a hand gripping; 3 studies tested a joystick movement; and 8 studies tested various upper limb tasks with 8 different interventions.

Language-related tasks were used in only one study. Finally, other studies did not fit into the 3 above mentioned categories. Eight studies performed laterality judgement tasks; 5 studies used MI tests and questionnaires; 2 tested neurofeedback; and 1 study test whole body MI.

Not all studies have evaluated patients with PD under the same conditions. Eleven studies evaluated patients in their off phase, 10 in the on phase, 6 in both phases and 14 did not mention this information.

#### 3.3.3 Outcomes for lower limb

Of these studies, 2 assessed walking in clinical conditions (40,41); 6 assessed brain activity with regional Cerebral Blood Flow (rCBF) using a Positron Emission Tomography (PET) scans (45,46) as well as using functional Magnetic Resonance Imaging (fMRI) (25,44–46); execution time was also used (7 studies) during different tasks (28,43–45,47–49).

#### 3.3.4 Outcomes for upper limb

In the thumb-opposition studies, Dominey et al. (50) evaluated the execution time for MI and ME. Avanzino et al. (51) evaluated the timing error rate. Cunnington et al. (52) performed this task under PET scan and compared the rCBF. Leiguarda et al. (53) evaluated the firing rate of the globus pallidus internus using microelectrode recording.

For hand gripping, muscle activation by electromyography (EMG) and monopolar local filed potentials were evaluated (41,54).

All joystick movement studies were done under PET scan (55–57). In addition, 2 of them evaluated the execution time (55,56).

For studies with varied upper limb tasks, the evaluations were also heterogeneous. The execution time was evaluated in 3 studies (39,40,58); KVIQ was assessed in one study (56); F-waves were assessed by EMG (59,60); the amplitude of motor evoked potential by transcranial magnetic stimulation (TMS) (60,61); movement-related potentials by electroencephalogram (62); and local field potentials by electrode recording (63).

#### 3.3.5 Outcomes for verbal task

Péran et al. (64) used the number of correct responses as well as fMRI as means of assessment.

#### 3.3.6 Outcomes for laterality judgment

Reaction time and error rate were measured for all these studies. MEP amplitude was assessed using TMS (65). fMRI was used in 2 studies (66,67).

#### 3.3.7 Outcomes for MI tests and questionnaire

Several tests were used in the different studies. The score of these studies was used as an outcome. There were the KVIQ, Motor Imagery Questionnaires (MIQ-R), Gait Imagery Questionnaire (GIQ), Chaotic Motor Imagery Assessment. The execution time was also measured for the BBT (29,68).

#### 3.3.8 Outcomes for neurofeedback intervention

In these non-RCTs studies, fMRI and UPDRS scores were used (69,70).

#### 3.3.9 Outcomes for MI of whole body

The rCBF was assessed using PET scan (71).

#### 3.3.10 Main results for lower limb (8 studies: 257 participants)

Firstly regarding execution time of walking in MI, 3 studies showed that there was no significant difference between PD/HS-MI (28,44,47). Cohen et al. (43) also found no significant difference between patients with PD with and without freezing of gait (FOG).

Secondly, still concerning execution time of walking but for PD/HS-ME, Peterson et al. (28) showed that patients with PD are slower than HS (p<0.001). It has been shown that patients with FOG were slower than patients without FOG in normal walking (p=0.03) as well as walking through a narrow doorway (p<0.0001) (43,44).

Maillet et al. (45) showed that patients with PD in off phase had significantly different durations during MI of walking (MI and ME data combined) compared to HS (p<0.03) while in the on phase (MI and ME data combined) there was no significant difference when compared to HS. Weiss et al. (46) assessed the difference between active and inactive transcranial stimulation in patients. When stimulation was active and for ME condition, patients walked 51% further (p<0.001), 57% faster (p<0.001) and took 30% longer steps (p<0.001).

Now regarding brain activity, Maillet et al. (45) showed that MI of walking in patients with PD compared to HS increased brain activation in premotor-parietal cortices and pontomesencephalic tegmentum and decreased brain activation in motor and frontal associative areas, basal ganglia, thalamus and cerebellum. Maidan et al. (49) found that patients with PD compared to HS had higher activation in the frontal, parietal, temporal, and occipital lobes during MI of usual walking (p<0.039). Huang et al. (48) showed that during MI of walking, patients with PD without FOG compared to controls had more brain activity in bilateral supplementary area, right superior temporal, and right medial superior frontal gyrus (p<0.041). Weiss et al. (46) showed that with or without deep brain stimulation in subthalamic nucleus, MI of walking induced activity in the supplementary motor area and the right superior parietal lobule against a rest condition (p<0.05). In terms of the difference in FOG, Snijders et al. (47) showed that FOG patients increased brain activity on fMRI in the mesencephalic locomotor region during MI of gait compared to non-FOG patients (p<0.05).

#### 3.3.11 Main results for thumb-opposition task (4 studies: 52 participants)

Dominey et al. study (50) showed that patients with PD were 69.8% slower compared to HS in execution time of thumb-opposition task (MI and ME data combined) (p<0.000). Avanzino et al. (48) found that when the task was performed in a 0.5 Hz timing and the auditory cue was removed, patients with PD made more errors when continuing the task in both MI (p=0.04) and ME (p=0.045) conditions, which was not the case for a 1.5 Hz timing. In Cunnington et al. study (52), the degree of activation in the supplementary motor area was normal in patients with PD when they were both in the “off” and “on” medication states during MI compared to rest (p<0.000).

#### 3.3.12 Main results for hand gripping task (2 studies: 32 participants)

Kobelt et al. (41) measured muscle activity by EMG and showed significant activation of the deltoideus pars clavicularis (p=0.001) and biceps brachii (p=0.007) during hand gripping task in MI as compared to a resting state. There was, however, no significant difference in activation between MI and rest in the extensor digitorum and flexor carpi radialis muscles. Fischer et al. (54) recorded local field potentials with TMS. They showed that beta activity decreased significantly for MI at the two highest force levels compared to rest (range: p<0.01–0.05) as well as for ME at all force levels (p<0.001); gamma activity increased significantly at MI at the two highest force levels again compared to rest (range: p<0.01–0.05) as well as for ME at all force levels (range: p<0.01–0.05).

#### 3.3.13 Main results for joystick movement (3 studies: 35 participants)

Thobois et al. (55) observed that patients with PD were slower with their more affected side than with the other side to perform the joystick movement task in both MI and ME conditions (range: 10.8–13.7%, p<0.05). In another study by Thobois et al. (56), were no significant difference in execution time between MI and ME. Samuel et al. (57) showed that when performing the task, patients with PD compared to HS had in MI a less activity in dorsolateral and mesial frontal cortex (p<0.01); and in ME a less activity in right dorsolateral frontal cortex and basal ganglia (p<0.01). The ability to retain movements previously made in MI as well as in ME was not different between PD and HS groups (57).

#### 3.3.14 Main results for varied upper limb tasks (8 studies: 265 participants)

Yágüez et al. (39) conducetd a pre-post clinical trial in patients with PD. They investigated writing movement and execution time to perform ideograms. The intervention was first a practice phase in MI and then a phase in ME. No difference was found for large ideograms between pre- and post-training in terms of movement duration and tangential velocity. However, a difference in execution time was found between baseline and post-ME practice sessions (p=0.014) as well as between post-MI and post-ME session with, in both cases an improvement after the ME practice phase (p=0.031).

Sabaté et al. (40) showed that sequential finger movements took 70% (p<0.001) longer in MI and 80% (p<0.001) longer in ME for patients with PD when compared to HS. Regarding the difference between MI and ME in patients with PD, Sabaté et al. (58) found a significant difference in favour of ME in execution time for a fast cyclic (p<0.001) and for a slow continuous movement tasks (p<0.001); but no significant difference for a slow cyclic movement task. Bek et al. (59) compared the KVIQ score before and after MI instructions and no significant changes were observed between patients with PD and HS. Gündüz et al. (60) measured F-waves during thumb abduction. They showed that the average amplitude of F-waves significantly increased during MI and ME compared to rest conditions in both patients with PD non-apraxia (P=0.005) and HS (P=0.028) groups. Tremblay et al. (61) measured the MEP amplitude of two hand muscles in the resting state and during the MI of a scissor cutting task. No significant change was detected between conditions in patients with PD while a significant difference was found in HS (p<0.05).

#### 3.3.15 Main results for verbal task (1 study: 10 participants)

Péran et al. (64) compared 3 tasks in patients with PD: *object naming*, *action word* related to the object and *mental simulation* of the action with the object. They found that in comparison to object naming, mental simulation demonstrated a greater level of activation in the prefrontal cortex bilaterally and in the parietal-occipital junction bilaterally (p<0.001).

#### 3.3.16 Main results for laterality judgment task (8 studies: 293 participants)

The laterality judgement tasks are all task is known to engage an implicit MI process. Four studies (50,72–74) divided the participants into groups according to the most affected side. Amick et al. (72) found that patients with PD right-sided symptoms group made more errors than HS in judging laterality (p=0.01), but the left-sided symptoms group did not show a significant difference in error rates compared to the HS group. The results of Conson et al. (73) showed that patients with PD had a higher reaction time to determine the laterality of a body that corresponded to their most affected side compared to the other side (range: p<0.006–0.028). However, no significant difference was found in terms of reaction time and accuracy between patients with right-sided symptoms and patients with left-sided symptoms (73). In the Dominey et al. (50), patients with PD were slower compared to HS to determine letter symmetry and hand laterality by 19.3% (p<0.0001). Scarpina et al. (74) and Helmich et al. (67), in a similar protocol, did not find significant difference in reaction time and accuracy among patients with PD with right-sided symptoms, patients with PD with left-sided symptoms and HS; between patients with PD with right-sided symptoms and HS; and between patients with PD with and without tremor and HS. Additionally, patients with PD with tremors demonstrated higher levels of imagery-related activity in the somatosensory area 3a when compared to both patients with PD without tremors and HS (p<0.01) (67).

#### 3.3.17 Main results for MI tests and questionnaire (5 studies: 232 participants)

Heremans et al. (29,68) found that patients with PD were slower on the BBT in MI and ME compared to HS (range: 16.7–30.4%; p<0.01–0.02). Regarding the influence of cues in BBT, there was no significant difference in execution time between MI with cues and ME, whereas MI without cues was slower than ME (p<0.05).

Several studies used MI tests and questionnaires. There was no significant difference between patients with PD and HS for the MIQ-R, KVIQ-20, CMIA, GIQ. Heremans et al. (68) and Peterson et al. (75) investigated KVIQ in patients with PD phase on, off, and HS and no significant was found among groups. For the GIQ no significant difference was found between patients with PD with FOG and without FOG (73).

Perspective preference during MI was assessed with KVIQ visual subscale (indicating clarity of the image). Patients with PD choose in 71.5% internal perspective (corresponds to a first-person view), in 26.3% external perspective (corresponds to a third-person view), in 0.4% both and in 2.3% no perspective could be chosen. Now with KVIQ kinaesthetic subscale (indicating intensity of the sensations), patients with PD choose in 73.3% internal, in 25.2% external, in 0.3% both and 1.4% no perspective could be chosen (41).

#### 3.3.18 Main results for neuro-feedback intervention (2 studies: 28 participants)

Tinaz et al. (69) used MI tasks that allowed positive neurofeedback activation. Subramanian et al. (70), showed a significant improvement of 37% (p=0.042) on the UPDRS score between pre- and post-intervention in the experimental group while the control group showed no significant difference. Tinaz et al. (69), showed no significant difference in patients with PD between pre- and post-intervention on the MDS-UDPRS-III score.

#### 3.3.19 Main results for MI of whole body (1 study: 22 participants)

Mori et al. (71) measured rCBF between patients with PD and HS during standing position. During MI, no significant difference was shown between groups. In contrast, during ME, patients with PD against HS showed a significant increase in the right cerebellar vermis and left paracentral gyrus, and a significant decrease in the bilateral middle frontal gyrus.

## 4 Discussion

Motor Imagery has been used in sport and performance activities and has attracted considerable interest since the 1980s (76). This technique has been adapted to PD patient’s rehabilitation with promising results, although there are still a small number of RCTs studies published (22–25, 31–38). Among the included studies (53 studies), there were few RCTs (12 studies) with an average PEDro score of 6.6, which can be considered as medium to high quality. The protocols as well as the outcomes measured were heterogeneous and there were no RCTs with specific outcomes for upper limbs or speech other than the UPDRS score. The population of RCTs and descriptive studies was quite young with a low severity level (i.e., H&Y score). Indeed, the most RCTs excluded patients with a score greater than 3. It is therefore not possible to conclude on the applicability of MI in patients with PD who have a higher severity. MI should therefore be used as early as possible before cognitive impairment prevents its use. Taking these aspects into account, the results should be treated with caution as methodological biases need to be resolved before conclusions can be drawn.

In complement of RCTs, we also investigated descriptive and non-RCTs to determine how MI have been used in the PD population. It is noted that for MI of walking, there was no difference between subjects with PD and HS, whereas, as expected, in ME patients with PD were slower than HS. This would suggest that patients with PD have relatively poor sensorimotor prediction capacities, as the tight relation between the duration of ME and MI (i.e. isochrony) seems broken (77). It is also found that patients with PD have similar scores to HS in MI questionnaires (such as KVIQ, MIQ-R, GIQ), which means that they can practice MI. The presence of cues (visual and auditory) was also found to improve the abilities of patients with PD in MI. There were various tasks for the upper limbs in this section including BBT, thumb opposition tasks which means that many tasks can be adapted and need to be investigated for the upper limb.

Collectively, it can therefore be said that MI of walking can be used along a corridor of varying distance with execution time as a measurement method. Walking speed as well as TUG can be interesting outcomes to be assessed at regular intervals to monitor progress. Then, for the upper limb, global and patient-specific tasks can be investigated by measuring the execution time. UPDRS score allows task monitoring for the upper and lower limb. Motor symptoms, assessed by the UPDRS, showed no significant difference between the two groups in RCTs. However, part 3 of the UPDRS includes items for both the upper and lower limbs and we have seen that the RCTs were targeted at the lower limbs. As MI protocol did not encompass all aspects evaluated in the UPDRS, this may explain the fact that there was no change (78).

Even though we did not set date limits we could not include many studies. Indeed, this is a recent topic of interest as, the first study included here was published in 1997 and, the first RCT included in this review dates from 2007. Among the studies excluded, 21 were ongoing clinical trials and whose results were not yet published, so we can see that there is an interest in this topic and that there will be more data in the next few years, which conditioning the update of this review in the next years.

Our contribution to this study was to guide and facilitate the use of MI in clinical practice, as well as to highlight the main results observed in these studies in terms of improvements in motor symptoms, balance, gait, and quality of life. Indeed, MI is a technique that does not require any equipment, it is easy and safe to set up and just requires a learning phase beforehand. In a context where the prevalence of PD is increasing it is important to empower patients and give them tools that they can use at home by completing other treatments.

The main limitation of this study was the fact that, for the descriptive and non-RCT studies, only the main tasks and outcomes of MI were analysed. Although the diversity of tasks and results were observed, we focused on the tasks with the best results, perhaps other interesting elements could be explored. Another limitation is that the most important studies included in this review (RTCs) excluded the most severe PD. Therefore, it is not known whether the recommendations raised here can be useful for more seriously ill patients.

Despite the limited number of RTCs focusing on MI in patients with PD, combined with diverse protocols, outcomes, and potential biases, the findings offer a promising outlook, especially in addressing walking and balance impairments. However, research on upper limb function or speech remains scant. Future studies in this area must involve larger participant cohorts and adopt more specific protocols tailored to the unique challenges posed by upper limb impairments. The criteria for assessing outcomes related to walking and balance align with recommendations from the French National Authority for Health, providing a valuable standard for evaluating MI interventions in PD.

In conclusion, it is crucial to acknowledge that this scoping review underscores the necessity for further research and updates in the coming years. The ongoing RCTs registered in clinical trial databases highlight the evolving landscape of MI interventions for PD, suggesting that a comprehensive and updated systematic review will be vital to capture the latest advancements and insights in this field.

## Author contributions

**MM, and ET**: Formal Analysis, Methodology, Project administration, Writing – original draft. **MB, EM, and NG**: Data curation, Funding acquisition, Methodology, Resources, Supervision, Writing – original draft. **AVS, and YS**: Resources, Visualization, Writing – original draft, Writing – review & editing.

## Funding

This work was supported by the ‘‘ANER’’ program, from Région Bourgogne Franche Comté (contract ANER PARK-IMAGE, 2021Y-08279).

## Conflict of interest

The authors declare that the research was conducted in the absence of any commercial or financial relationships that could be construed as a potential conflict of interest.

## Data Availability

The original contributions presented in this study are included in this article/supplementary material, further inquiries can be directed to the corresponding authors.

## Supplementary Material

## Notes

### Competing Interest Statement

The authors have declared no competing interest.

### Funding Statement

This work was supported by the ANER program, from Region Bourgogne Franche Comte (contract ANER PARK-IMAGE, 2021Y-08279).

